# *PCSK9* genetic variants and risk of vascular and non-vascular diseases in Chinese and UK populations

**DOI:** 10.1101/2023.08.15.23294117

**Authors:** Michael V Holmes, Christiana Kartsonaki, Ruth Boxall, Kuang Lin, Nicola Reeve, Canqing Yu, Jun Lv, Derrick A Bennett, Michael R Hill, Ling Yang, Yiping Chen, Huaidong Du, Iain Turnbull, Rory Collins, Robert J Clarke, Martin D Tobin, Liming Li, Iona Y Millwood, Zhengming Chen, Robin G Walters, China Kadoorie Biobank Collaborative Group

## Abstract

**Background:** Lowering low-density lipoprotein cholesterol (LDL-C) through PCSK9 inhibition represents a new therapeutic approach to preventing and treating cardiovascular disease (CVD). Phenome-wide analyses of *PCSK9* genetic variants in large prospective biobanks can help to identify unexpected effects of PCSK9 inhibition.

**Methods:** The prospective China Kadoorie Biobank genotyped >100,000 participants with follow-up for fatal and non-fatal disease events. A *PCSK9* genetic score (*PCSK9*-GS) was constructed using three single nucleotide polypmorphisms at the *PCSK9* locus associated with directly-measured LDL-C, including a loss-of-function variant (rs151193009). Logistic regression gave estimated odds ratios (ORs) for *PCSK9*-GS associations with CVD and non-CVD outcomes, scaled to 1-standard deviation (SD) lower LDL-C.

**Results:** *PCSK9*-GS was associated with lower apolipoprotein B, and with lower risks of carotid plaque (n=8340 cases; OR=0.61 [95%CI: 0.45-0.83]; P=0.0015), major occlusive vascular events (n=15,752; 0.80 [0.67-0.95]; P=0.011), and ischaemic stroke (n=11,467; 0.80 [0.66-0.98]; P=0.029). However, *PCSK9*-GS was also associated with higher risk of hospitalisation with chronic obstructive pulmonary disease (COPD: n=6836; 1.38 [1.08-1.76]; P=0.0089), and with even higher risk of fatal exacerbations among individuals with pre-existing COPD (n=730; 3.61 [1.71-7.60]; P=7.3×10^−4^). Risk of acute upper respiratory tract infection (URTI) was also increased (n=1,095; 2.18 [1.34-3.53]; P=0.0016), as previously reported in UK Biobank, with a pooled OR after meta-analysis of 1.87 ([1.38-2.54]; P=5.4×10^−5^). We also replicated a previously-reported association with self-reported asthma (pooled OR 1.17 ([1.04-1.30]; P=0.0071). There was heterogeneity between the associations of *PCSK9*-GS and a polygenic LDL-C score for each of COPD hospitalisation (P-het=0.015), fatal COPD exacerbation (P-het=0.0012), and URTI (P-het=0.013).

**Conclusions:** LDL-C-lowering *PCSK9* genetic variants are associated with lower risk of subclinical and clinical atherosclerotic vascular disease, but higher risks of respiratory diseases which appear unrelated to LDL-C. Pharmacovigilance studies may be required to monitor patients treated with therapeutic PCSK9 inhibitors for exacerbations of respiratory diseases or respiratory tract infections.

## Background

Lowering of low-density lipoprotein cholesterol (LDL-C) is an established, efficacious approach for treatment and prevention of occlusive cardiovascular disease (CVD)^1^. While statins represent the main class of LDL-C lowering drug used in routine clinical practice, new drugs targeting different components of LDL-C metabolism have gained clinical traction, including inhibition of Niemann-Pick C1-Like 1 (i.e. ezetimibe^2^) and of proprotein convertase subtilisin kinase 9 (PCSK9)^3, 4^.

Compared with statins, PCSK9 inhibitors (e.g. evolocumab, inclusiran) are relatively new, with much less comprehensive long-term data concerning their benefits and potential side-effects. Two large randomised trials have reported that inhibition of PCSK9 over a period of around 2 to 3 years significantly reduced the risk of major CVD events in high-risk patients with prior history of vascular disease^3, 4^. Moreover, these trials have demonstrated a reduction in atheroma volume^5^ and that the reduction in CVD events was proportional to the reduction in LDL-C achieved^6^. However, there was suggestive evidence of excess risk of diabetes associated with PCSK9 inhibition^7^. As with statins^8^, confirmation (or refutation) of any excess risk of diabetes will likely require meta-analysis of multiple large trials with patients treated and followed up for longer durations. Little is known about the long-term effects of PCSK9 inhibition on a wide range of other major disease outcomes.

Human genetics can be used to predict the likely effects of therapeutic modification of a drug target on disease outcomes^9–11^. Several well-established examples have either predicted or recapitulated findings from randomised clinical trials of lipid lowering (such as HMGCR^12^ and CETP^13^ inhibitors) and other therapies (including inhibitors of interleukin 6 receptor^14, 15^, secretory phospholipase A2-IIA^16^, and lipoprotein-associated phospholipase A2^17, 18^). This approach has provided evidence corroborating findings from randomised clinical trials that PCSK9 inhibition leads to reduced LDL-C^19^, with a corresponding reduction in risk of CVD^20, 21^. Similarly, genetic studies provide strong evidence that, as with statins, LDL-C lowering by PCSK9 inhibition may increase the risk of diabetes^20, 22–24^. In addition to diabetes, analyses in UK Biobank (UKB) of a functional variant of *PCSK9* (rs11591147) suggested possible excess risks of several non-vascular disease outcomes, including asthma and respiratory tract infections^22,25^, although these associations were only nominally significant and require replication.

In this study we sought to further investigate the potential impact of therapeutic PCSK9 inhibition. In approximately 100,000 genotyped individuals from the prospective China Kadoorie Biobank (CKB), we assessed the associations of a *PCSK9* genetic score (*PCSK9*-GS) with lipids, lipoproteins and a range of vascular and non-vascular disease outcomes, including (where available) those previously reported in UKB. Where applicable, we further undertook meta-analyses of CKB and UKB data for respiratory disease and other relevant disease outcomes.

## Methods

### Study design and population

CKB is a prospective cohort study of 512,713 adults aged 30-79 years, recruited between 2004 and 2008 from 5 urban and 5 rural areas across China^26^. In brief, baseline information was collected via a laptop-based questionnaire (including demographic and lifestyle factors and medical history) and physical measurements (including anthropometry, blood pressure, and lung function). Duplicate lung function measurements were conducted using a portable handheld ‘Micro spirometer’ (Micro Medical Limited, Rochester, Kent, England), as previously described^27^, from which forced expiratory volume in 1s (FEV1) and forced vital capacity (FVC) were derived; the distributions of values for FEV1/FVC, with large numbers of individuals with values of 1, indicated systematic errors in data collection in two recruitment regions (Qingdao and Haikou, see **Supplementary Figure 1**), so participants in these regions were excluded from analyses requiring spirometry data. A non-fasting blood sample was collected (with time since last meal recorded) and separated into plasma and buffy-coat fractions for long-term storage. Resurveys of random 5% subsets of the cohort were conducted at periodic intervals, and the second resurvey in 2013-2014 included measurements of carotid intima media thickness (CIMT) and plaque^28^.

Incident disease outcomes were identified from long-term follow up through electronic linkage of each participant’s unique national identification number to the Chinese national health insurance system, and to established regional registries for death and major diseases (cancer, ischaemic heart disease, stroke, and diabetes). Health insurance records included detailed information about each hospital admission (e.g. disease description, International Statistical Classification of Diseases and Related Health Problems, 10th Revision [ICD-10] code, and procedure or examination codes). All reported cases of disease outcomes from different sources were centrally checked, reviewed and standardised by clinicians.

### Biomarker assays

Plasma concentrations of total cholesterol, LDL-C, high-density lipoprotein cholesterol (HDL-C), triglycerides, apolipoprotein B (ApoB), and apolipoprotein A-I from baseline samples were quantified in 18,181 CKB participants using clinical chemistry assays at the Wolfson Laboratory (AU 680 clinical chemistry analysers, Beckman Coulter [UK] Ltd., Wycombe, United Kingdom) using manufacturers’ reagents, calibrators, and settings^29^. In a subset of 4,442 individuals, a high-throughput targeted ^1^H-NMR metabolomics platform^30, 31^ was used to generate spectra from which 225 lipid and other metabolic measures were simultaneously quantified by Nightingale Health Ltd. (Helsinki, Finland; previously known as Brainshake Ltd).

### GWAS genotyping

Genome-wide genotyping data were available for a subset of 100,706 CKB participants, comprising ~30,000 participants selected for nested case-control studies of CVD and respiratory disease, and ~70,000 being randomly-selected from the remaining participants^32^. Region-specific principal component analysis identified 6,107 individuals with ancestry not local to the region in which they were recruited (of whom 774 had clinical biochemistry measurements), who were excluded from region-stratified analyses. To avoid biases due to over-representation of disease cases in the genotyped dataset, we constructed a subset of 70,914 individuals representative of the full CKB cohort in which such over-representation was eliminated; this was a preliminary version of the population-representative sample described elsewhere^32^.

### Instrument selection and derivation of weighted PCSK9 locus score

In 17,687 samples with both genotyping and LDL-C data, LDL-C was regressed on age, age^2^, sex, study area, fasting time, and fasting time^2^. The residuals were rank inverse-normal transformed (RINT) and genome wide association analysis of the transformed variable was conducted using BOLT-LMM v2.3.2 ^33^. Summary statistics for variants within a 1Mbp window around the *PCSK9* structural gene were evaluated using FINEMAP v1.1^34^, identifying the model with the highest posterior probability of explaining association at the locus with LDL-C, comprising a set of three single nucleotide polypmorphisms (SNPs; rs151193009, rs2495477, rs11206517: log_10_ Bayes Factor=47.6). These were used to construct a weighted genetic score, with SNP weights corresponding to the beta coefficients for association of the SNPs with RINT LDL-C. To account for linkage disequilibrium between the SNPs, the per-allele beta coefficients were derived from a region-stratified, multivariable (mutually adjusted) model including dosages for the 3 SNPs, age, age^2^, sex, study area, fasting time, fasting time^2^, and region-specific principal components, with exclusion of 774 individuals with non-local ancestry (**Supplementary Table 1**), which were applied to individuals without LDL-C data. To avoid potential bias from using internally-derived weights, beta coefficients for participants with LDL-C data were instead derived using 100-fold block-jack-knifing, from regressions that excluded random 1% subsets of participants to whom those beta coefficients were assigned^35^. For each individual with genotyping data, the sum of SNP dosages, weighted by their corresponding beta coefficients, gave a *PCSK9* gene score calibrated to the predicted effect on LDL-C.

### Derivation of weighted LDL-C genetic score

An LDL-C genetic risk score was derived using SNPs that were previously identified as independently-associated with LDL-C (at P<5×10^−8^) in GWAS of Europeans in the Global Lipids Genetics Consortium (GLGC)^36^. Of 76 independent association signals, 2 SNPs were monomorphic in CKB; 64 of the remaining 74 SNPs showed an association with LDL-C in CKB that was consistent with that in GLGC (i.e. directionally consistent or no significant effect size heterogeneity at P<0.05/74). These 64 SNPs were used to construct an LDL-C GS in CKB (further described in the **Supplementary Methods** and **Supplementary Table 2**), allowing a comparison of disease associations arising from LDL-C lowering by *PCSK9* variants with those from LDL-C lowering overall. This LDL-C score did not include variants at the *PCSK9* locus.

### Disease endpoints

The detailed vascular and non-vascular disease outcomes used in the present study and their corresponding ICD-10 codes are provided in **Supplementary Table 3**. Vascular disease outcomes included major coronary events (MCE: non-fatal myocardial infarction, fatal ischaemic heart disease, or coronary revascularisation); fatal/nonfatal ischaemic stroke; fatal/nonfatal intracerebral haemorrhage; major occlusive vascular events (MOVE: consisting of fatal/nonfatal major coronary events or ischaemic stroke); fatal CVD; and major vascular events (MVE: fatal/nonfatal myocardial infarction, coronary revascularisation procedures, stroke, or fatal CVD). These analyses used a common set of controls that excluded all individuals who self-reported prior CHD, stroke or transient ischaemic attack at baseline, or who experienced any form of MVE during the follow-up period.

The main non-vascular outcomes were organised by anatomical site and consisted of: combined incident, self-reported, and screen-detected cases of diabetes; and incident events of each of chronic obstructive pulmonary disease (COPD); chronic kidney disease; chronic liver disease; eye disease; malignancy; and non-vascular mortality (see **Supplementary Table 3**). Controls for these diseases excluded individuals with self-reported history of that disease at baseline (where available). Phenome-wide analysis was conducted in the population subset using incident disease outcomes in the ICD-10 code range A00 to N99 grouped together (defined in **Supplementary Table 4**), with no exclusions for prevalent disease for either cases or controls.

To dissect the principal incident COPD outcomes, which comprised a mixture of incident and recurrent disease, we defined prevalent COPD cases at baseline as being those with FEV1/FVC less than the lower limit of normal as predicted from the Global Lung Function 2012 spirometry reference equations^37^ according to participants’ ancestry, age, height, and sex (**Supplementary Figure 2**). Amongst those with spirometry-defined prevalent COPD at baseline (where available), we further defined those experiencing one or more incident COPD outcomes as exacerbations of COPD. Moreover, we further analysed COPD outcomes according to disease severity, according to whether incident COPD cases or cases of COPD exacerbation were fatal (COPD identified as cause of death) or non-fatal. These analyses excluded all participants in the 2 recruitment regions with systematic errors in spirometry data collection. Analyses of upper respiratory tract infections (URTI) excluded cases occurring before 2009, to avoid biases due to a spike in reported cases (>10% of all CKB cases across all years) in Zhejiang in 2007-2008.

### Statistical analysis

For continuous variables measured in the CVD nested case-control subset, such as blood biochemistry and NMR metabolomics, data transformation and analyses were stratified by recruitment region: following linear regression of each variable on age, age^2^, sex, case ascertainment category, and up to nine region-specific genetic principal components, the residuals underwent stratum-specific RINT. For CIMT, which was measured at the second resurvey in a randomly-selected subset of surviving participants, adjustments and data transformation were as above except without adjustment for case ascertainment. Associations of the weighted *PCSK9*-GS with these continuous traits were assessed by linear regression within each region with inverse-variance-weighted fixed-effect (IVW-FE) meta-analysis of the resulting estimates. SNP associations with spirometry were extracted from previous GWAS in CKB of FEV1/FVC^38^ with IVW-FE meta-analysis of the resulting estimates. LDL-C variance explained and F-statistics for the *PCSK9* and LDL-C genetic scores were determined by comparison of linear models including or excluding the scores, using the R anova() function.

Disease outcome associations with *PCSK9*-GS were assessed by logistic regression within each region with adjustment for sex, age, age^2^ and up to nine region-specific genetic principal components, and estimates were combined using IVW-FE meta-analysis. For all disease outcomes, cases and controls were selected from the population-representative subset of 70,914 genotyped individuals, plus additional cases identified among the remaining genotyped individuals. However, to avoid potential ascertainment bias, additional cases were excluded from the analysis if they were selected for genotyping on the basis of a different disease outcome that might separately be associated with altered *PCSK9* expression (see **Supplementary Table 5**).

For confirmation of the therapeutic effects of inhibition of PCSK9 in reducing risk of CVD outcomes^4, 39^, we assessed a combination of simple and composite outcomes, with a Bonferroni-adjusted threshold of significance (P<0.05/3=0.0167) based on 3 independent tests (determined by spectral decomposition of the correlation matrix for these related endpoints^40^). Similarly, we used a Bonferroni-adjusted threshold of significance (P<0.05/(7 + 3 prior tests) = 0.005) for non-vascular disease associations (type 2 diabetes, COPD, chronic kidney disease, chronic liver disease, malignant neoplasms, eye disease, and non-vascular mortality), and also for the phenome-wide scan (P<0.05/(41 + 10 prior tests) = 9.8×10^−4^). For sensitivity analyses of subgroups, analyses were performed in the full dataset with additional adjustment for recruitment region.

For disease outcomes previously reported as nominally associated with a functional *PCSK9* variant in a recent study of UKB participants by Rao et al^22^, associations of *PCSK9*-GS with seven (out of 12) diseases that were available in CKB were investigated as follows. We used identical case/control definitions (or the closest possible match), based on ICD-10 codes or responses to the baseline questionnaire (**Supplementary Table 6**); for cerebrovascular disease, the analysis was limited strictly to the population-representative subset – this was to minimise risk of bias due to over- or under-representation of particular disease subtypes, since the case-control part of the genotyped dataset is not representative of the full spectrum of cerebrovascular disease. Replication of these prior associations was assessed at 5% false discovery rate (FDR, Benjamini-Hochberg). Heterogeneity was assessed using Cochran’s Q-statistic with a Bonferroni-adjusted significance threshold (P<0.05/7).

In order to compare effect estimates, these were scaled to the same difference in LDL-C (1-SD lower) as follows: the UKB study by Rao et al.^22^ (and Nelson et al.^25^) used a missense variant in *PCSK9* (rs11591147) to quantify the associations with risk of diseases. We took the per-allele rs11591147 LDL-C estimate from the Global Lipids Genetics Consortium^41^ (per-allele beta coefficient: 0.497 SD) and scaled the disease association estimates reported by Rao et al^22^ so that they were equivalent to 1-SD lower LDL-C, as used for the CKB effect estimates. The scaled estimates derived from UKB and CKB were combined in IVW-FE meta-analyses, and Cochran’s Q statistic was used to assess heterogeneity.

All analyses used release version 15 of the CKB database and were performed using SAS software (version 9.3; SAS Institute, Inc) or R v4.2.1.

## Results

### Participant characteristics

In the population-representative subset of 70,914 genotyped participants, the mean age at study baseline was 52.1 (SD 10.7), 59.7% were female, and 45.8% were from urban regions. Stratifying individuals according to whether they were carriers of a *PCSK9* functional variant (see below), no appreciable differences between these two groups were observed at baseline for a wide range of CVD risk factors (**Table 1**), including height, adiposity, blood pressure, physical activity, alcohol drinking, and smoking. Similar proportions of the two groups had prior diagnoses of CVD, hypertension, or diabetes, or were taking statins.

**Table 1:** Baseline characteristics of the study population, dichotomised as non-carriers/carriers of the loss-of-function *PCSK9* variant rs151193009-T.

| Characteristics | rs151193009 genotype |  |
| --- | --- | --- |
|  | CC<br>(N=69,088) | CT/TT<br>(N=1,826) |
| Age (years), mean (SD) | 52.1 (10.7) | 52.5 (10.9) |
| Female, % | 59.7 | 61.2 |
| Urban region, % | 45.8 | 47.2 |
| LDL-C (mmol/L), mean (SD) | 2.36 (0.69) | 1.99 (0.69) |
| Height (cm), mean (SD) | 158.8 (8.3) | 159.1 (8.2) |
| BMI (kg/m<sup>2</sup>), mean (SD) | 23.8 (3.4) | 24.0 (3.5) |
| Waist/hip ratio, mean (SD) | 0.88 (0.07) | 0.88 (0.07) |
| SBP (mmHg), mean (SD) | 131.5 (21.4) | 132.5 (21.8) |
| DBP (mmHg), mean (SD) | 78.0 (11.2) | 78.8 (11.5) |
| Physical activity (MET h/d), mean (SD) | 20.6 (14.0) | 20.6 (14.3) |
| High school or higher education, % | 21.0 | 21.8 |
| Household income ≥20,000 yuan, % | 40.1 | 40.4 |
| Current regular alcohol drinking, % | 15.5 | 15.5 |
| Current regular smoker, % | 25.9 | 24.2 |
| Stroke/TIA<sup>a</sup>, % | 1.7 | 1.7 |
| Coronary heart disease<sup>a</sup>, % | 3.6 | 4.2 |
| Hypertension, % | 11.6 | 12.9 |
| Diabetes<sup>b</sup>, % | 6.1 | 6.2 |
| Taking statins, % | 1.9 | 1.6 |

BMI: body mass index; SBP: systolic blood pressure; DBP: diastolic blood pressure; MET: metabolic equivalent of task
<sup>a</sup>self-reported physician diagnosed. <sup>b</sup>self-reported physician diagnosed or screen-detected at baseline

### Characterisation of the PCSK9 genetic score

In 17,687 participants with both genotyping and LDL-C data, three SNPs in/around *PCSK9* (rs151193009, rs2495477 and rs11206517) were independently associated with LDL-C at genome-wide significance (P<5×10^−8^) in a multivariable model, with per-allele effects on LDL-C of 0.65 SD (SE=0.05), 0.10 SD (SE=0.01), and 0.16 SD (SE=0.03), respectively (**Supplementary Table 1**). The SNP with the strongest effect, rs151193009C>T, is a known loss-of-function variant that disrupts binding of the PCSK9 protein prodomain to heparan sulphate proteoglycans (HSPGs), an essential step in PCSK9-mediated degradation of LDL receptors (LDLR)^42^. When combined into the weighted *PCSK9*-GS, the three SNPs explained 1.2% of the variance of LDL-C (F-statistic=206), and showed a consistent association with LDL-C between women and men, and across geographical regions, age groups, and smoking or alcohol behaviour categories (**Supplementary Figure 3**). Scaled to a 1-SD lowering of LDL-C (equivalent to 27.4 mg/dl), *PCSK9*-GS was associated with lower concentrations of ApoB (−0.95 SDs, 95%CI: −1.08, −0.82; P=6.7×10^−46^) and, more weakly, a lowering of Lp(a) (−0.14 [−0.27, −0.01]; P=0.038). No significant associations were detected with triglycerides (−0.01 [−0.14, 0.12]; P=0.87) or HDL-C (−0.10 [−0.23, 0.03]; P=0.13) (**Figure 1**). Using detailed measures of lipoproteins and lipids quantified by NMR-spectroscopy, we identified associations of *PCSK9*-GS with lower concentrations of cholesterol among the smaller subclasses of very low-density lipoproteins and across intermediate- and low- density lipoprotein subclasses (**Supplementary Figure 4**). After accounting for multiple testing, there was no significant association with inflammation biomarkers including glycoprotein acetyls, C-reactive protein, or fibrinogen, nor were associations identified with other non-lipid blood biochemistry measures (**Supplementary Figures 4-5**).

**Figure 1:**
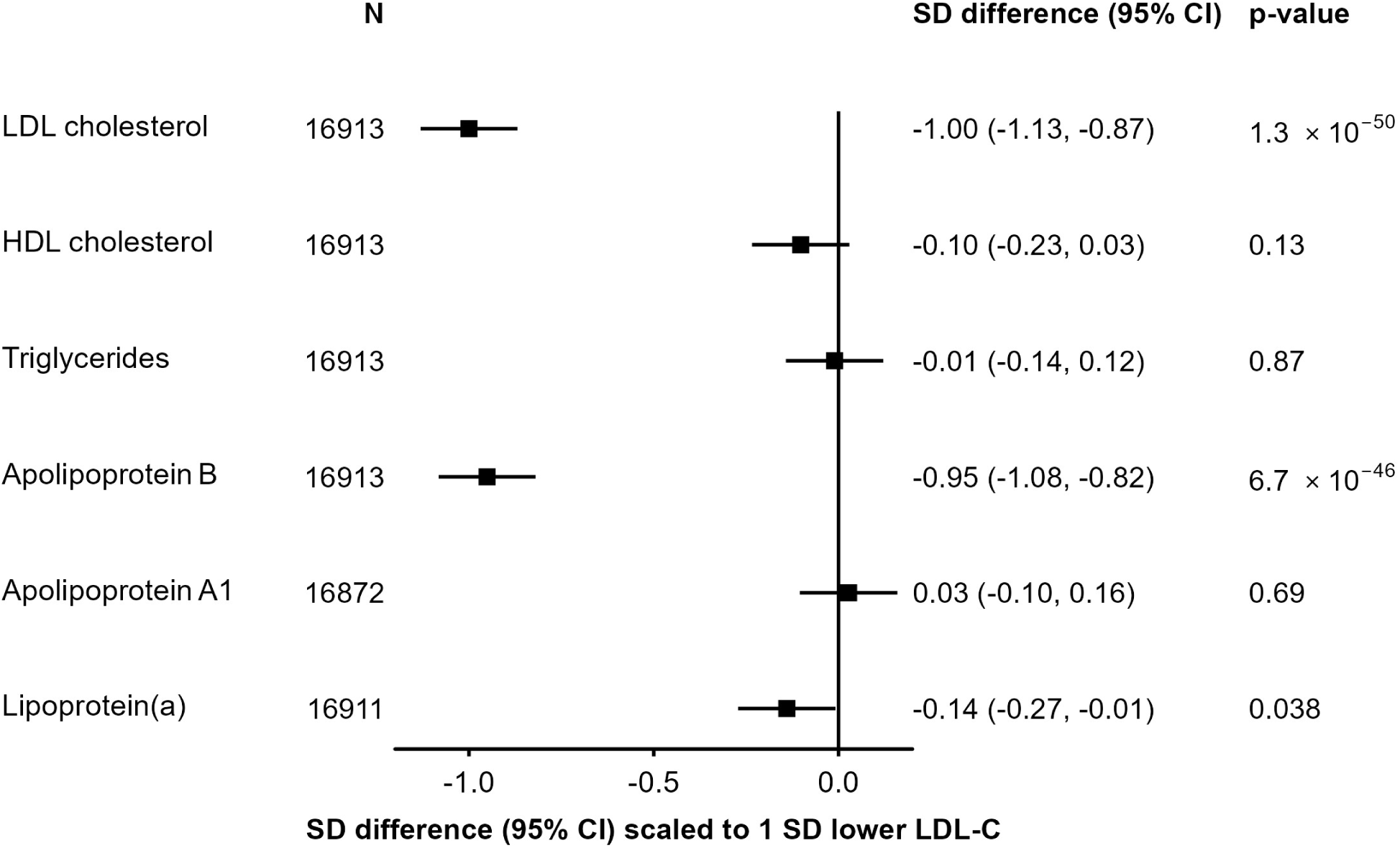
Associations of *PCSK9* genetic score with major blood lipids, and apolipoproteins. Estimates are standardized beta coefficients from linear regressions of the weighted *PCSK9* genetic score, scaled to a 1-SD lowering of LDL-cholesterol, calculated as an inverse-variance-weighted average of region-specific estimates with adjustment for age, age^2^, sex, ascertainment, and region-specific principal components.

### Association of PCSK9 with subclinical and clinical vascular outcomes

As expected, *PCSK9*-GS was associated with clinical CVD outcomes. Scaled to 1 SD lower LDL-C, there was reduced risk of MOVE which was significant after adjustment for multiple testing (n=15,752; OR=0.80 [0.67, 0.95]; P=0.011) and nominally significant reduced risk of ischaemic stroke (n=11,467; 0.80 [0.66, 0.98]; P=0.029); for MCE, MVE, and fatal CVD, the associations were directionally consistent but not statistically significant (**Figure 2A**). In addition, the score was associated with lower CIMT (n=20,896; −0.24 SDs [−0.36, −0.12]; P=0.00012) and 37% lower odds of carotid plaque (n=8340 cases; 0.61 [0.45, 0.83]; P=0.0015) (**Supplementary Table 7**). Conversely, no association of *PCSK9*-GS was observed with risk of intracerebral haemorrhage (n=5,906; 1.04 [0.81, 1.34]; P=0.74).

**Figure 2:**
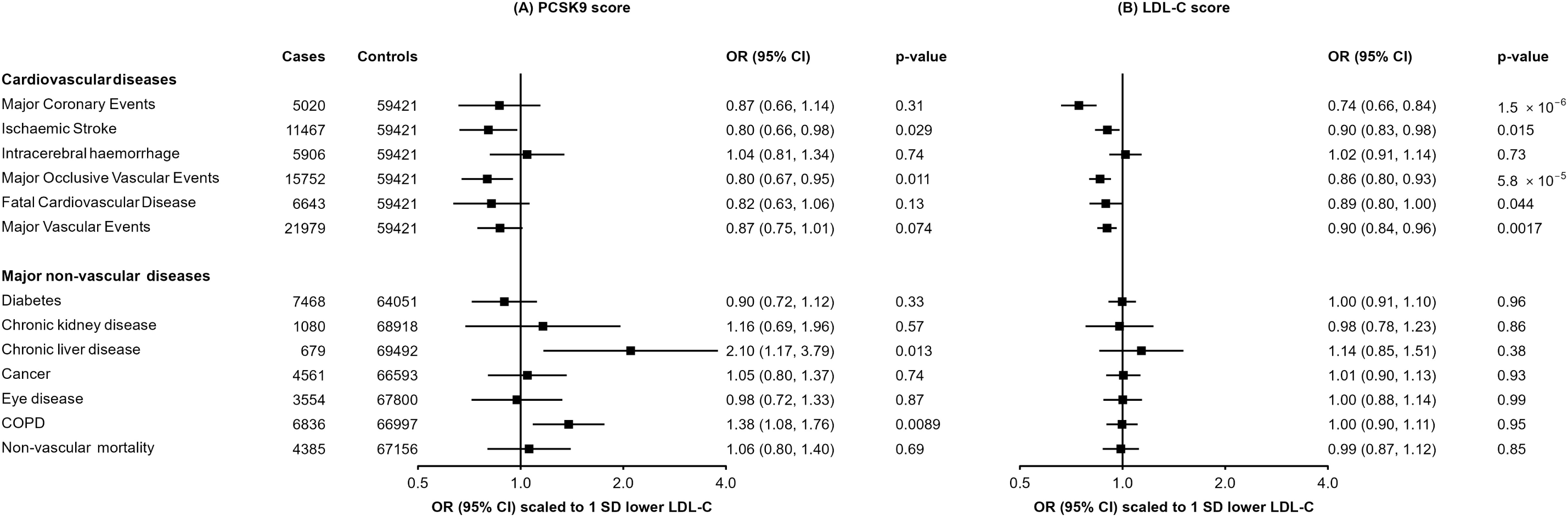
Associations of genetic scores for (A) *PCSK9* and (B) LDL cholesterol with risk of vascular and non-vascular disease endpoints. Odds ratios are estimated using logistic regression of the weighted *PCSK9* genetic score, scaled to a 1-SD lowering of LDL cholesterol, calculated as an inverse-variance-weighted average of region-specific estimates with adjustment for age, age^2^, sex, ascertainment, and region-specific principal components.

This pattern of associations with risk of vascular disease was very similar to that obtained when using a genetic score for LDL-C (**Figure 2B**) constructed with 64 SNPs from across the genome but which did not include SNPs at the *PCSK9* locus (variance explained=5.6%, F-statistic=1002; **Supplementary Table 2**). For example, using the LDL-C genetic score, 1 SD lower LDL-C was associated with lower risk of MCE (0.74 [0.66, 0.84]; P=1.5×10^−6^) and ischaemic stroke (0.90 [0.83, 0.98]; P=0.015), but not intracerebral haemorrhage (1.02 [0.91, 1.14]; P=0.73).

### Association of genetic scores with major non-vascular outcomes

We further assessed the associations of the *PCSK9* and LDL-C genetic scores with major non-vascular outcomes. No associations were identified for *PCSK9*-GS with diabetes, chronic kidney diseases, eye diseases, malignant neoplasms, or non-vascular mortality (**Figure 2A**). There was a nominally significant positive association of higher LDL-C with risk of liver disease (2.10 [1.17, 3.79]; P=0.013), and a strongly suggestive positive association with risk of incident COPD (1.38 [1.08, 1.76]; P=8.9×10^−3^), although these were not significant after accounting for multiple testing. The LDL-C genetic score showed no association with any of these outcomes (**Figure 2B**). Association of *PCSK9*-GS was also investigated for a wide range of other disease outcomes available in CKB using a phenome-wide approach (**Supplementary Figure 6),** with no association meeting the pre-specified significance threshold.

### Replication of reported PCSK9 associations

For seven traits previously reported as being nominally-associated with a missense *PCSK9* variant (rs11591147) in UKB^22^, we tested association with *PCSK9*-GS in CKB (**Figure 3**). Scaled to a 1-SD lowering of LDL-C, association in CKB of *PCSK9*-GS with higher risk of URTI (n=1,095 cases; 2.18 [1.34, 3.35; P=1.6×10^−3^) was significant at 5% FDR and was consistent with the association observed in UKB (n=2,364 cases; 1.70 [1.15, 2.51]), providing an estimate after meta-analysis of 1.87 [1.38, 2.54] (P=5.4×10^−5^). A nominally significant association was also identified for self-reported doctor-diagnosed asthma (n=427 cases; 2.28 [1.12, 4.64]; P=0.02), again consistent with the association reported in UK Biobank (n=39,269 cases; 1.15 [1.02, 1.28]), with an estimate after meta-analysis of OR=1.17 [1.04, 1.30] (P=0.007). Although low case numbers limited power for replication in CKB, effect size estimates were consistent with those from UKB for the other disease outcomes, with the exception of breast cancer (P-het = 0.005).

**Figure 3.**
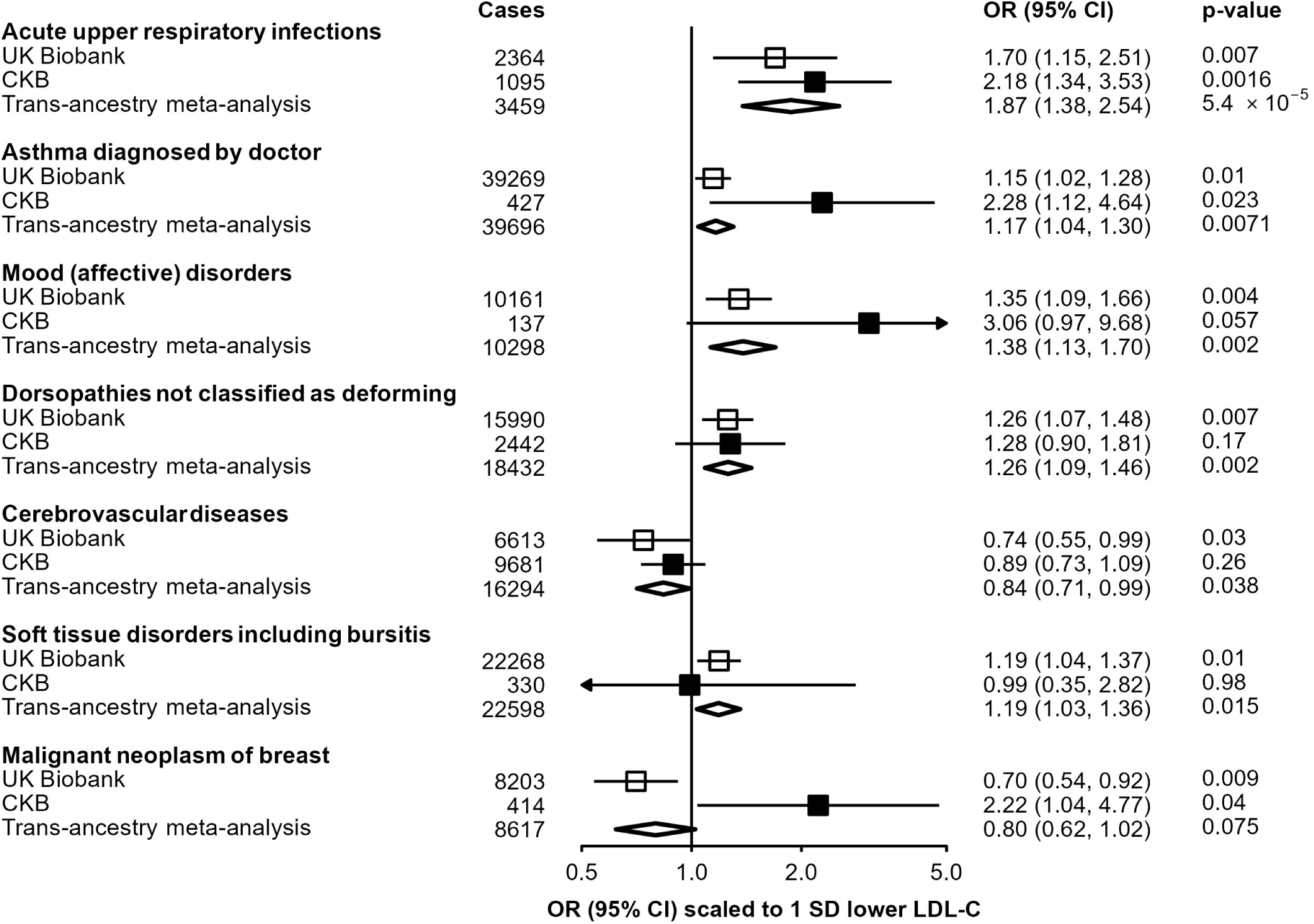
Replication in CKB of previously reported associations in UKB of *PCSK9* genetic scores with disease outcomes. Estimates in UKB (open boxes) originate from data published by Rao et al^22^, and scaled to a corresponding 1-SD lowering of LDL cholesterol. Odds ratios in CKB (filled boxes) are estimated using logistic regression of the weighted *PCSK9* genetic score, scaled to a 1-SD lowering of LDL cholesterol, calculated as an inverse-variance-weighted average of region-specific estimates with adjustment for age, age^2^, sex, ascertainment, and region-specific principal components. Trans-ancestry estimates (diamonds) are derived from inverse-variance-weighted fixed-effect meta-analysis.

### Association of PCSK9 genetic score with further respiratory endpoints

The association of *PCSK9*-GS with multiple respiratory-related diseases, in both UKB and CKB, was further investigated in related outcomes in CKB (**Figure 4**). The primary incident COPD outcome, reflecting either death or hospital admission due to COPD, was consistently observed across fatal (n=1632; 1.47 [0.93, 2.34]; P=0.098) or non-fatal (n=5204; 1.38 [1.05,1.80], P=0.019) events. However, there was no association of the genetic score with prevalent COPD, as defined by spirometry measurements at baseline (n=5,105; 0.96 [0.73, 1.26]; P=0.75), nor with baseline FEV1/FVC as a continuous measure (**Supplementary Table 8**). By contrast, the association with higher risk of an incident COPD event was stronger in those with pre-existing COPD (2212 exacerbations among 6.409 individuals with spirometry-defined COPD; 1.91 [1.12, 3.24]; P=0.017). The increased risk was most clearly observed in those for whom the COPD exacerbation was fatal (730 deaths due to acute exacerbations of COPD: 3.61 [1.71, 7.60]; P=7.1×10^−4^), although this was not significantly different from risk of a non-fatal exacerbation (1,482 non-fatal exacerbations: 1.58 [0.86, 2.91]; P=0.14; P-het=0.24). The association of *PCSK9*-GS with URTI persisted after excluding individuals with spirometry-defined COPD and no differences were found across geographical regions, age groups, sex, smoking status, and alcohol consumption (**Supplementary Figure 7**).

**Figure 4.**
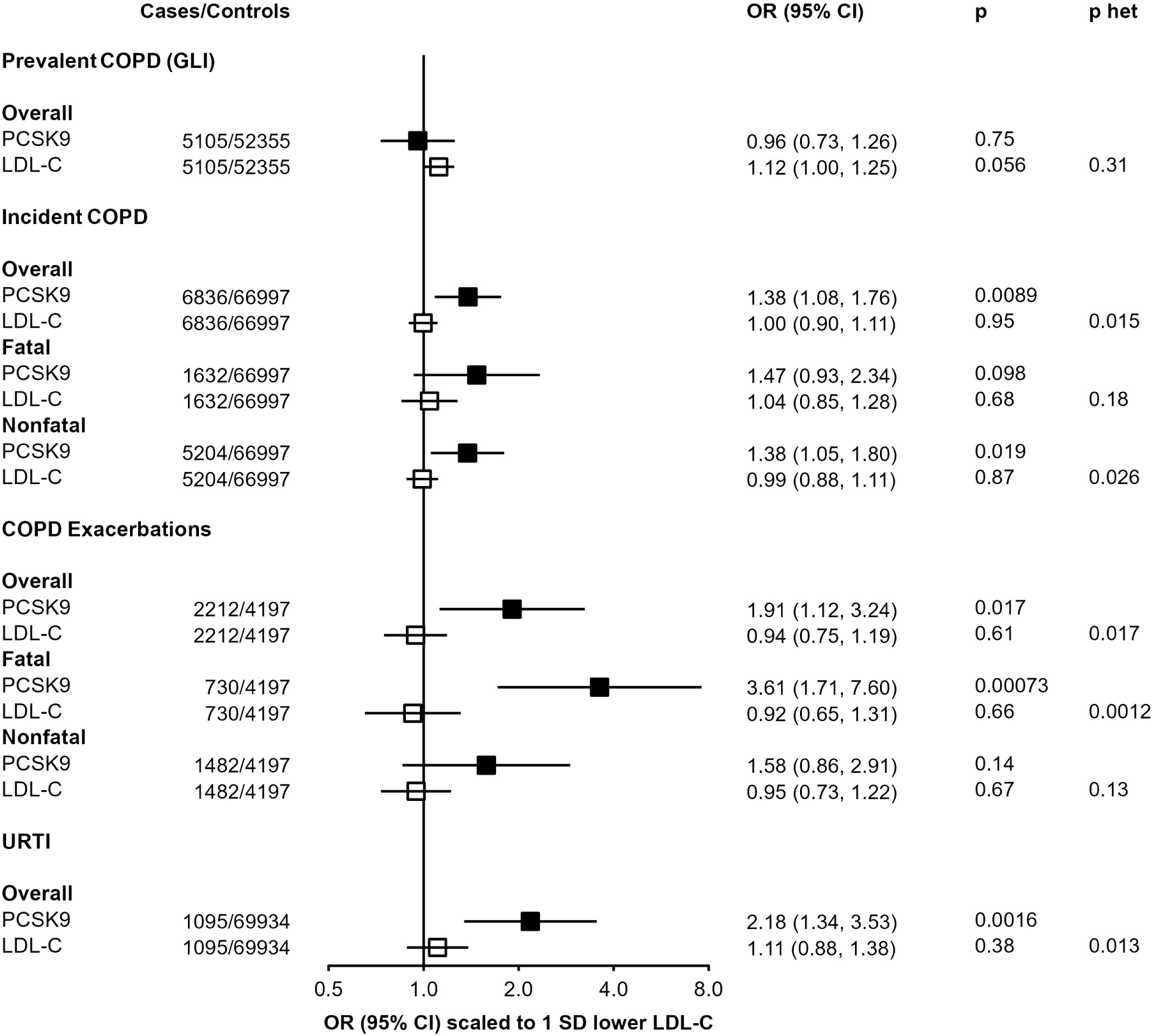
Associations of *PCSK9* and LDL-C genetic scores with respiratory disease endpoints. Odds ratios are estimated using logistic regression of the weighted *PCSK9* genetic score, scaled to a 1-SD lowering of LDL cholesterol, calculated as an inverse-variance-weighted average of region-specific estimates with adjustment for age, age^2^, sex, ascertainment, and region-specific principal components. Prevalent COPD defined on basis of spirometry (see Methods). COPD exacerbations are incident events occurring in individuals with spirometry-defined prevalent COPD. P het is the p-value from a test for heterogeneity for estimates of PCSK9 and LDL-C genetic instruments.

The association with risk of COPD exacerbation in CKB was not replicated in UKB using a different functional *PCSK9* variant (**Supplementary Table 9**), although power was limited due to a much smaller number of recorded cases (468 in UKB vs 2212 cases in CKB), and it is unclear to what extent the aetiology or definition of acute exacerbation of COPD differed between the two biobanks which derive from very different populations with contrasting exposures to other risk factors for respiratory disease^43^. After meta-analysis, the pooled OR was 1.45 [0.91, 2.32] (P=0.12), but with significant heterogeneity between the 2 estimates (P-het = 0.032).

By contrast with *PCSK9*-GS, the LDL-C GS shows no association with any of these respiratory disease outcomes (**Figure 4**). When scaled to the same predicted difference in LDL-C, there was significant heterogeneity between associations with *PCSK9*-GS and LDL-C GS for both URTI (*PCSK9*: 2.18 [1.34, 3.53]; LDL-C GS: 1.11 [0.88, 1.38]; P-het=0.013) and fatal COPD exacerbations (*PCSK9*: 3.61 [1.71, 7.60]; LDL-C GS: 0.92 [0.75, 1.19]; P-het=0.0012).

## Discussion

We used genetic variants, including a loss-of-function variant, to mimic pharmacological inhibition of PCSK9 to gain insight into expected therapeutic effects on a broad range of disease outcomes in a Chinese population. As expected, genetic inhibition of PCSK9 was associated with lower levels of LDL-C and ApoB, less subclinical atherosclerosis, lower risk of MOVE, including ischaemic stroke, and a directionally concordant effect on risk of MCE. While these associations are confirmatory of previously reported results in European ancestry populations, as the relationship of PCSK9 inhibition with risk of vascular disease is well documented from large-scale cardiovascular outcome trials^4, 39^ and other genetic studies^20, 22, 44, 45^, they demonstrate that PCSK9 inhibition is likely also to be effective for the treatment of CVD in East Asian populations, in whom mean LDL-C levels are lower than in Western populations. Conversely, the present study also indicated that PCSK9 inhibition is potentially associated with higher risks of URTI, acute exacerbation of COPD, and asthma.

MR studies in Europeans have shown that a 1-SD reduction in LDL-C (equivalent to ~1 mmol/L) leads to a reduction in risk of CHD of approximately 40%^46^. This compares to a more modest association in CKB for a comparable outcome of around 25% per 1-SD lower LDL-C, although this is at least partly attributable to a smaller SD for LDL-C in CKB (0.69 mmol/L) than in European populations. When scaled to the same mmol/L reduction in LDL-C as in Europeans, the reduction in risk in CKB is 34%, which suggests consistency of effect across ancestral groups once appropriate comparisons are made^1^. A pre-specified secondary analysis of the FOURIER trial identified that treatment with a PCSK9 inhibitor led to a monotonic relationship between LDL-C lowering and risk of vascular events^6^. Similarly, the GLAGOV trial illustrated a dose response relationship between LDL-C lowering and volume of atherosclerosis^5^. Thus, although the population studied here had much lower LDL-C levels at baseline as compared to European studies (e.g. 2.35 (0.69 SD) mmol/L CKB vs 3.57 (0.87) UKB^46^), triangulation of our genetic findings with studies in Europeans and RCTs of LDL-C-lowering therapies, including PCSK9 inhibitors, supports the clinical efficacy of PCSK9 inhibition on vascular disease outcomes in patients of East Asian ancestry. Moreover, these findings provide further support for intensive LDL-C lowering therapy in high risk individuals (e.g. adding PCSK-9 inhibitor to statin) in order to achieve maximal beneficial effects.

In addition to confirming the expected associations with various forms of occlusive CVD, we further identified that *PCSK9*-GS, orientated to a lowering of LDL-C, was associated with a higher risk of URTI, acute exacerbations of COPD, and self-reported asthma. For URTI and asthma, our study findings replicate nominally significant findings in a previous phenome scan of another functional variant in *PCSK9* conducted within the UKB^22,25^. In addition to these three respiratory disease outcomes, meta-analysis of our results with those from UKB strengthened evidence for an association of *PCSK9* genetic variants with mood disorders and with non-deforming dorsopathies (diseases of the spine). Previous studies have identified association of *PCSK9* with severe forms of dorsopathy^47^ which we were unable to investigate in CKB, a population-based study of middle-aged adults.

The findings of the present study linking PCSK9 inhibition with lung infections have biological plausibility. The mechanism by which PCSK9 influences LDL-C is by mediating lysosomal degradation of LDLR, so that inhibition of PCSK9 leads to higher levels of LDLR in the liver^48^ and a concomitant reduction in blood LDL-C levels. However, PCSK9 binds to and mediates degradation of not only LDLR but also very low-density lipoprotein receptors (VLDLR)^49, 50^. PCSK9, LDLR and VLDLR are each also expressed in the lung^51^, where VLDLR and LDLR are involved in the initiation of infection by several classes of respiratory viruses (e.g. human rhinovirus, some types of coronavirus), which bind to VLDLR and LDLR on the cell surface, and are internalised via receptor-mediated endocytosis^52, 53^. Inhibition of the LDL receptor in cultured human tracheal epithelial cells lowers the infectivity of human rhinovirus^54^. Thus, it is possible that the relationship of genetic variants mimicking therapeutic inhibition of PCSK9 with a higher risk of URTI (which can underlie acute exacerbations of COPD^55^, and worsen asthma symptoms^56^) may arise from elevated levels of VLDLR or LDLR in lung tissue. Since the proposed mechanism is via higher susceptibility to respiratory viruses, leading to an increase in exacerbations of COPD (which are often infectious in nature), we would not expect an association with prevalent COPD, as observed – the pathophysiology of COPD development is itself separate, a potential example of disease incidence vs progression being related to discrete aetiological pathways.

A review article in 2013 suggested that dual PCSK9 and statin therapy might amplify potential safety issues, including those related to respiratory infections, given that both act to increase levels of LDLR and VLDLR^57^. What is unclear is whether PCSK9 inhibitors in clinical use (or under development) alter pulmonary VLDL receptor levels and, thus, whether genetic findings reflecting life-long systemic changes in PCSK9 have clinical relevance to patients receiving long-term treatment with tissue-localised PCSK9 inhibitors. While various parameters determine the tissue distribution of monoclonal antibodies^58^, current evidence suggests that evolocumab has limited tissue distribution^59, 60^. Furthermore, inclisiran, a small interfering RNA inhibitor of *PCSK9* that is currently under development^61^, features N-acetylgalactosamine conjugation that renders it hepatotropic^62^. Consequently, limited tissue penetration of biological agents may mean that the genetic associations we identify do not have major clinical relevance.

The available evidence indicates that neither of two large phase III trials led to any excess risk of respiratory disease in individuals treated with PCSK9 inhibition for a mean ~2 to 3 years^4, 39^. However, the association of *PCSK9*-GS with exacerbations of COPD may indicate that PCSK9 inhibition may confer higher risk only in individuals with existing respiratory disease, perhaps triggered by URTI. Clinical trials may not have included many such participants, thus limiting their power to detect any excess risk within the short period of trial treatment. While some phase II trials did show a non-significant excess risk of URTI in those treated with evolocumab^63–65^, such flu-like symptoms may be common following treatment with monoclonal antibodies in general^66^ and, as such, it is not clear whether these findings reported in RCTs represent a reaction to the treatment modality as opposed to PCSK9 inhibition itself.

Despite the consistency of associations of genetic variants in *PCSK9* and risk of lung diseases in these two large biobanks, and the underlying biological plausibility, there are several other factors to consider in interpreting these findings. First, the genetic findings may be a false positive. For example, we failed, albeit based on a small number of cases and using a different functional *PCSK9* variant, to replicate the association observed in CKB with a similar COPD exacerbation endpoint in UKB. In the HUNT biobank in Norway, no association of *PCSK9* genetic variants with risks of asthma (6858 cases) or COPD (6685 cases) was identified, although there was a nominal association with rhinitis^67^; in that study, the association with URTI was not reported and it is also unclear whether COPD was prevalent or represented acute exacerbations of disease. Second, direct comparisons between different populations are complicated by functional differences between the genetic variants available in different ancestries. The variant with the strongest effect in the CKB *PCSK9* score modifies the PCSK9 HSPG-binding domain, directly impacting a key step in LDL receptor degradation^42^. By contrast, the UK Biobank functional variant (rs11591147) instead exerts its effect by modulating local protein structure^68^. While both lead to lower LDL-C, suggesting overall similarities in their biological consequences, their relative impact on differing PCSK9-mediated pathways may vary. Third, the association of *PCSK9*-GS with URTI might arise due to the life-long effects of genetic perturbations that alter human biology^69^, meaning that treatment with a PCSK9 inhibitor at a specific time (e.g. in later life) need not lead to a similarly altered risk of lung disease. Fourth, as discussed above, limited tissue penetration of biological therapeutics may mean that VLDLR and LDLR levels in lung parenchyma are unaltered. Fifth, even if the genetic associations with respiratory disease foretell findings in humans from inhibition of PCSK9 that have yet to be identified, the relevance of the magnitude of these effects and how to frame these in terms of absolute risk are unclear. While there is a strong basis for calibration of genetic estimates for vascular disease risk of LDL-C lowering variants to the equivalent from a therapeutic trial^70^, no such calibration exists for non-vascular disease outcomes such as respiratory disease, which poses a translational challenge.

This study demonstrates the advantages of leveraging large-scale genetic data from prospective biobanks in diverse populations to inform drug target validation, repurposing, and design and conduct of randomised trials. Apart from providing improvements in statistical power, a key advantage is in accessing the considerable heterogeneity in the genetic and environmental characteristics of different populations, to inform the utility and application of therapies in varying contexts globally. Further, our study used functional variants that are present at appreciable frequency only in one ancestry, which, when findings are consistent across different functional variants in different ancestral groups with very different environmental exposures, reinforces the credibility of the data. However, despite this, our findings remain hypothesis generating and require further validation in future studies involving much larger numbers of well-characterised disease outcomes.

## Conclusion

This study provides evidence that genetic variants in *PCSK9* that lower LDL-C and risk of CVD are also associated with higher risks of respiratory disease, in particular acute URTI and acute exacerbations of COPD. While further genetic and clinical evidence is needed to confirm (or refute) these findings, they raise the possibility that, in individuals with pre-existing COPD, a careful risk-benefit evaluation of LDL-C lowering through inhibition of PCSK9 may be required. Pharmacovigilance may be warranted to ascertain whether these findings have relevance to patients being treated with PCSK9 inhibitors.

## Declarations

### Ethics approval

Ethical approval for CKB was obtained jointly from the University of Oxford, the Chinese Centre for Disease Control and Prevention (CCDC) and the regional CCDC from the 10 study areas. All participants provided written informed consent.

### Data availability

The China Kadoorie Biobank (CKB) is a global resource for the investigation of lifestyle, environmental, blood biochemical and genetic factors as determinants of common diseases. The CKB study group is committed to making the cohort data available to the scientific community in China, the UK and worldwide to advance knowledge about the causes, prevention and treatment of disease. For detailed information on what data is currently available to open access users and how to apply for it, visit: https://www.ckbiobank.org/data-access.

Researchers who are interested in obtaining raw data from the China Kadoorie Biobank study that underlies this paper should contact. A research proposal will be requested to ensure that any analysis is performed by bona fide researchers and - where data is not currently available to open access researchers - is restricted to the topic covered in this paper. Sharing of genotyping data is currently constrained by the Administrative Regulations on Human Genetic Resources of the People’s Republic of China. Access to these and certain other data is available only through collaboration with CKB researchers.

### Author contributions

Data collection and analysis – MVH, CK, RB, KL, NR, CY, JL, DAB, MRH, LY, YC, HD, IT, MDT, IYM, RGW; funding –ZC, LL, RGW, IYM, RJC, MRH, RC; study oversight: MVH, RGW; manuscript draft and revision – MVH, RGW, IYM, ZC. All authors read and approved the final manuscript.

## Acknowledgements

The most important acknowledgement is to the participants in the study and the members of the survey teams in each of the 10 regional centres. We also thank the project development and management teams based at Beijing, Oxford, and the 10 regional centres. China’s National Health Insurance provides electronic linkage to all hospital treatments.

## License

This research was funded in whole, or in part, by the Wellcome Trust [212946/Z/18/Z, 202922/Z/16/Z, 104085/Z/14/Z, 088158/Z/09/Z]. For the purpose of Open Access, the author has applied a CC-BY public copyright licence to any Author Accepted Manuscript version arising from this submission.

