## Supplementary information for "*PCSK9* genetic variants and risk of vascular and non-vascular diseases in Chinese and UK populations"

#### Members of the China Kadoorie Biobank collaborative group:

**International Steering Committee:** Junshi Chen, Zhengming Chen (PI), Robert Clarke, Rory Collins, Yu Guo, Liming Li (PI), Chen Wang, Jun Lv, Richard Peto, Robin Walters.

**International Co-ordinating Centre, Oxford:** Daniel Avery, Derrick Bennett, Ruth Boxall, Sushila Burgess, Ka Hung Chan, Yiping Chen, Zhengming Chen, Johnathan Clarke; Robert Clarke, Huaidong Du, Ahmed Edris, Hannah Fry, Simon Gilbert, Mike Hill, Pek Kei Im, Andri Iona, Maria Kakkoura, Christiana Kartsonaki, Hubert Lam, Kuang Lin, Mohsen Mazidi, Iona Millwood, Sam Morris, Qunhua Nie, Alfred Pozarickij, Paul Ryder, Saredo Said, Dan Schmidt, Paul Sherliker, Becky Stevens, Iain Turnbull, Robin Walters, Baihan Wang, Lin Wang, Neil Wright, Ling Yang, Xiaoming Yang, Pang Yao.

**National Co-ordinating Centre, Beijing:** Xiao Han, Can Hou, Qingmei Xia, Chao Liu, Jun Lv, Pei Pei, Canqing Yu.

**Regional Co-ordinating Centres:**

**Gansu:** Gansu Provincial CDC – Caixia Dong, Pengfei Ge, Xiaolan Ren. Maiji CDC – Zhongxiao Li, Enke Mao, Tao Wang, Hui Zhang, Xi Zhang. **Haikou:** Hainan Provincial CDC – Jinyan Chen, Ximin Hu, Xiaohuan Wang. Meilan CDC – Zhendong Guo, Huimei Li, Yilei Li, Min Weng, Shukuan Wu. **Harbin:** Heilongjiang Provincial CDC – Shichun Yan, Mingyuan Zou, Xue Zhou. Nangang CDC – Ziyan Guo, Quan Kang, Yanjie Li, Bo Yu, Qinai Xu. **Henan:** Henan Provincial CDC – Liang Chang, Lei Fan, Shixian Feng, Ding Zhang, Gang Zhou. Huixian CDC – Yulian Gao, Tianyou He, Pan He, Chen Hu, Huarong Sun, Xukui Zhang. **Hunan:** Hunan Provincial CDC – Biyun Chen, Zhongxi Fu, Yuelong Huang, Huilin Liu, Qiaohua Xu, Li Yin. Liuyang CDC – Huajun Long, Xin Xu, Hao Zhang, Libo Zhang. **Liuzhou:** Guangxi Provincial CDC – Naying Chen, Duo Liu, Zhenzhu Tang. Liuzhou CDC – Ningyu Chen, Qilian Jiang, Jian Lan, Mingqiang Li, Yun Liu, Fanwen Meng, Jinhuai Meng, Rong Pan, Yulu Qin, Ping Wang, Sisi Wang, Liuping Wei, Liyuan Zhou. **Qingdao:** Qingdao CDC – Liang Cheng, Ranran Du, Ruqin Gao, Feifei Li, Shanpeng Li, Yongmei Liu, Feng Ning, Zengchang Pang, Xiaohui Sun, Xiaocao Tian, Shaojie Wang, Yaoming Zhai, Hua Zhang, Licang CDC – Wei Hou, Silu Lv, Junzheng Wang. **Sichuan:** Sichuan Provincial CDC – Xiaofang Chen, Xianping Wu, Ningmei Zhang, Weiwei Zhou. Pengzhou CDC – Xiaofang Chen, Jianguo Li, Jiaqiu Liu, Guojin Luo, Qiang Sun, Xunfu Zhong. **Suzhou:** Jiangsu Provincial CDC – Jian Su, Ran Tao, Ming Wu, Jie Yang, Jinyi Zhou, Yonglin Zhou. Suzhou CDC – Yihe Hu, Yujie Hua, Jianrong Jin Fang Liu, Jingchao Liu, Yan Lu, Liangcai Ma, Aiyu Tang, Jun Zhang. **Zhejiang**: Zhejiang Provincial CDC – Weiwei Gong, Ruying Hu, Hao Wang, Meng Wang, Min Yu. Tongxiang CDC – Lingli Chen, Qijun Gu, Dongxia Pan, Chunmei Wang, Kaixu Xie, Xiaoyi Zhang.

### Supplementary Methods

#### *Derivation of the LDL cholesterol genetic risk score*

The LDL cholesterol (LDL-C) genetic risk score (GS) was constructed by using single nucleotide polymorphisms (SNPs) previously identified as associated with LDL-C at $P<5x10^{-8}$ by the Global Lipid Genetics Consortium (GLGC)[36]. Prior to inclusion of each SNP in the LDL-C GS, we assessed heterogeneity of the per-allele LDL-C effect estimate between CKB and GLGC using Cochran's Q statistic; SNPs without evidence of heterogeneity between CKB and GLGC (after Bonferroni multiple testing correction for 74 SNPs) were weighted using GLGC association estimates. SNPs with effect sizes in CKB significantly different from the estimate from GLGC were excluded from the instrument if (i) they did not show an association with LDL-C in CKB (at P<0.05/74) or (ii) if the direction of association was opposite to that reported in GLGC. The association of the weighted LDL-C GS with disease outcomes was assessed by logistic regression, stratified by region, and with adjustment for age, $age^2$, sex and 2-9 regional genetic principal components.

#### *Replication of COPD exacerbation in UK Biobank*

To replicate the association of *PCSK9* variants with COPD exacerbations, we obtained estimates of the per-allele estimate of a functional *PCSK9* variant (rs11591147) with risk of COPD exacerbations defined as the presence of one or more acute exacerbations from HES data among 25,030 participants in the UK Biobank with COPD GOLD 2-4 (defined as FEV1/FVC<0.7 and percent predicted FEV1<80%) at study baseline.  The estimate was scaled to an equivalent per-1-SD lower LDL-C using the effect estimate from GLGC, and was compared with our estimate from CKB. UK Biobank analyses were performed under project approval 648.

### Supplementary Tables

**Supplementary Table 1:** Association of SNPs included in the PCSK9 genetic score with LDL-cholesterol in univariate and multivariate analysis.

| | | | Univariate Model | | | | Multivariate Model | | | |
|---|---|---|---|---|---|---|---|---|---|---|
| SNP | Effect/ other allele | Effect allele freq. | N | effect (SD) | SE | P-value | N | effect (SD) | SE | P-value |
| rs151193009 | C/T | 0.986 | 16913 | 0.630 | 0.047 | 3.0E-41 | 16913 | 0.646 | 0.047 | 3.2E-43 |
| rs2495477 | A/G | 0.738 | 16913 | 0.058 | 0.013 | 1.0E-05 | 16913 | 0.104 | 0.014 | 1.2E-13 |
| rs11206517 | G/T | 0.060 | 16913 | 0.102 | 0.023 | 9.9E-06 | 16913 | 0.159 | 0.025 | 8.7E-11 |

Analyses were stratified by region and combined by inverse-variance-weighted fixed effect meta-analysis. LDL-cholesterol measures from clinical biochemistry assays from each region were adjusted for age, $age^2$, fasting time, fasting $time^2$, ascertainment group, sex, and 2-9 regional principal components, and rank inverse normal transformed. Effect allele is defined as the LDL-C increasing allele.

**Supplementary Table 2:** Associations with LDL-cholesterol in GLGC and CKB of 74 previously identified LDL-cholesterol-associated SNPs

| | | | | | | CKB Univariate Model | | | | GLGC (As Used for Score) | | | | | |
|---|---|---|---|---|---|---|---|---|---|---|---|---|---|---|---|
| rsID | Chr:Pos | Genes | Effect/ other allele | EAF (CKB) | EAF (1KG) | N | Estimate (SD) | SE | P-value | N | Estimate (SD) | SE | P-value | Het P-value | Included in CKB LDL-C GS? |
| rs10903129 | 1:25641524 | TMEM57 | G/A | 0.300 | 0.227 | 16913 | 0.035 | 0.012 | 0.003 | 169920 | 0.033 | 0.004 | 4.0E-19 | 0.885 | Yes |
| rs4587594 | 1:62906518 | DOCK7 | G/A | 0.812 | 0.823 | 16913 | 0.019 | 0.014 | 0.175 | 173007 | 0.049 | 0.004 | 3.0E-37 | 0.034 | Yes |
| rs6603981 | 1:92766395 | EVI5 | T/C | 0.960 | 0.975 | 16913 | 0.067 | 0.029 | 0.019 | 173056 | 0.034 | 0.004 | 2.0E-14 | 0.252 | Yes |
| rs646776 | 1:109620053 | *CELSR2/MYBPHL/PSRC1/SARS/SORT1 | T/C | 0.941 | 0.954 | 16913 | 0.213 | 0.023 | 9.6E-21 | 173021 | 0.160 | 0.004 | 1.0E-292 | 0.022 | Yes |
| rs267733 | 1:149225460 | ANXA9 | A/G | 0.964 | 0.974 | 16913 | -0.030 | 0.028 | 0.296 | 164562 | 0.033 | 0.005 | 5.0E-10 | 0.030 | Yes |
| rs2642438 | 1:219036651 | MOSC1 | G/A | 0.829 | 0.768 | 16913 | 0.065 | 0.015 | 1.2E-05 | 165470 | 0.035 | 0.004 | 4.0E-17 | 0.053 | Yes |
| rs903319 | 1:219052434 | MOSC1 | C/T | 0.297 | 0.299 | 16913 | 0.012 | 0.012 | 0.326 | 172998 | 0.027 | 0.004 | 8.0E-11 | 0.244 | Yes |
| rs2587534 | 1:232915962 | CR596412 | A/G | 0.761 | 0.769 | 16913 | 0.060 | 0.013 | 5.0E-06 | 172966 | 0.039 | 0.004 | 3.0E-26 | 0.130 | Yes |
| rs1367117 | 2:21117405 | APOB | A/G | 0.126 | 0.115 | 16913 | 0.099 | 0.016 | 1.9E-09 | 173007 | 0.119 | 0.004 | 2.0E-196 | 0.236 | Yes |
| rs3817588 | 2:27584716 | GCKR | T/C | 0.668 | 0.657 | 16913 | 0.034 | 0.012 | 0.004 | 172990 | 0.026 | 0.005 | 3.0E-08 | 0.511 | Yes |
| rs6544713 | 2:43927385 | ABCG8 | T/C | 0.006 | 0.005 | 16913 | -0.095 | 0.071 | 0.183 | 172940 | 0.081 | 0.004 | 6.0E-85 | 0.014 | Yes |
| rs4148218 | 2:43953086 | ABCG8 | G/A | 0.885 | 0.885 | 16913 | -0.005 | 0.017 | 0.762 | 173032 | 0.044 | 0.005 | 3.0E-21 | 0.005 | Yes |
| rs2710642 | 2:63003061 | EHBP1 | A/G | 0.688 | 0.730 | 16913 | 0.026 | 0.012 | 0.028 | 172994 | 0.024 | 0.004 | 3.0E-10 | 0.895 | Yes |
| rs2030746 | 2:121025958 | None | T/C | 0.469 | 0.496 | 16913 | 0.002 | 0.011 | 0.873 | 173024 | 0.021 | 0.004 | 2.0E-08 | 0.089 | Yes |
| rs16831243 | 2:135478814 | YSK4 | T/C | 0.407 | 0.385 | 16913 | 0.003 | 0.011 | 0.809 | 162945 | 0.038 | 0.006 | 8.0E-12 | 0.005 | Yes |
| rs2287623 | 2:169538401 | ABCB11 | G/A | 0.254 | 0.260 | 16913 | 0.033 | 0.012 | 0.008 | 170077 | 0.022 | 0.004 | 7.0E-09 | 0.392 | Yes |
| rs1250229 | 2:216012629 | *FN1 | C/T | 0.932 | 0.923 | 16913 | 0.025 | 0.023 | 0.278 | 173032 | 0.024 | 0.004 | 8.0E-09 | 0.982 | Yes |
| rs11563251 | 2:234344123 | UGT1A1/UGT1A10/UGT1A3/UGT1A4/UGT1A5/UGT1A6/UGT1A7/UGT1A8/UGT1A9 | T/C | 0.085 | 0.118 | 16913 | 0.012 | 0.020 | 0.557 | 172855 | 0.035 | 0.006 | 2.0E-08 | 0.280 | Yes |
| rs9875338 | 3:12271469 | *GSTM1L/PPARG | G/A | 0.918 | 0.931 | 16913 | 0.037 | 0.020 | 0.060 | 172895 | 0.027 | 0.004 | 3.0E-13 | 0.613 | Yes |
| rs17345563 | 3:133691893 | DNAJC13 | A/G | 0.898 | 0.882 | 16913 | 0.014 | 0.018 | 0.430 | 173048 | 0.036 | 0.006 | 3.0E-10 | 0.262 | Yes |
| rs7703051 | 5:74661243 | *COL4A3BP/HMGCR | A/C | 0.528 | 0.510 | 16913 | 0.078 | 0.011 | 7.2E-13 | 173015 | 0.073 | 0.004 | 5.0E-85 | 0.636 | Yes |
| rs6882076 | 5:156322875 | *TIMD4 | C/T | 0.733 | 0.727 | 16913 | 0.055 | 0.012 | 8.0E-06 | 173006 | 0.046 | 0.004 | 5.0E-33 | 0.469 | Yes |
| rs2294261 | 6:16217142 | *MYLIP | A/C | 0.097 | 0.083 | 16913 | 0.009 | 0.018 | 0.640 | 171831 | 0.033 | 0.004 | 5.0E-19 | 0.191 | Yes |
| rs1800562 | 6:26201120 | HFE | G/A | 0.999 | 1.000 | 10871 | -0.012 | 0.177 | 0.947 | 171209 | 0.062 | 0.008 | 2.0E-14 | 0.679 | Yes |
| rs2247056 | 6:31373469 | HLA-B/HLA-B*0707 | C/T | 0.853 | 0.834 | 16913 | -0.010 | 0.015 | 0.491 | 169288 | 0.025 | 0.004 | 6.0E-09 | 0.026 | Yes |
| rs868943 | 6:116444196 | FRK | G/A | 0.953 | 0.955 | 16913 | -0.005 | 0.025 | 0.837 | 170111 | 0.026 | 0.004 | 1.0E-12 | 0.218 | Yes |

| | | | | | | CKB Univariate Model | | | | GLGC (As Used for Score) | | | | | |
|---|---|---|---|---|---|---|---|---|---|---|---|---|---|---|---|
| rsID | Chr:Pos | Genes | Effect/ other allele | EAF (CKB) | EAF (1KG) | N | Estimate (SD) | SE | P-value | N | Estimate (SD) | SE | P-value | Het P-value | Included in CKB LDL-C GS? |
| rs1564348 | 6:160498850 | SLC22A1 | C/T | 0.004 | 0.004 | 16335 | 0.100 | 0.086 | 0.242 | 172989 | 0.048 | 0.005 | 3.0E-22 | 0.543 | Yes |
| rs12670798 | 7:21573877 | DNAH11 | C/T | 0.491 | 0.502 | 16913 | 0.015 | 0.011 | 0.175 | 173013 | 0.034 | 0.004 | 7.0E-16 | 0.092 | Yes |
| rs4722551 | 7:25958351 | None | C/T | 0.029 | 0.019 | 16913 | 0.036 | 0.030 | 0.239 | 172946 | 0.039 | 0.005 | 7.0E-16 | 0.916 | Yes |
| rs217386 | 7:44567220 | *DDX21/DDX56/DQ574505/NPC1L1/OGDH/TMED4 | G/A | 0.968 | 0.976 | 16913 | 0.017 | 0.030 | 0.559 | 173021 | 0.036 | 0.004 | 8.0E-22 | 0.529 | Yes |
| rs4240624 | 8:9221641 | AK055863 | A/G | 0.988 | 0.988 | 16913 | 0.146 | 0.051 | 0.004 | 171657 | 0.067 | 0.006 | 7.0E-27 | 0.122 | Yes |
| rs10102164 | 8:55584167 | *SOX17 | A/G | 0.214 | 0.207 | 16913 | 0.036 | 0.013 | 0.007 | 173037 | 0.032 | 0.005 | 3.0E-12 | 0.775 | Yes |
| rs2326077 | 8:59548473 | *CYP7A1/UBXN2B | C/T | 0.205 | 0.244 | 16913 | 0.041 | 0.014 | 0.002 | 172996 | 0.034 | 0.004 | 2.0E-19 | 0.619 | Yes |
| rs2737252 | 8:116733072 | TRPS1 | G/A | 0.699 | 0.703 | 16913 | 0.035 | 0.012 | 0.003 | 172950 | 0.031 | 0.004 | 1.0E-14 | 0.757 | Yes |
| rs2954022 | 8:126551803 | *TRIB1 | C/A | 0.430 | 0.445 | 16913 | 0.050 | 0.011 | 4.7E-06 | 172992 | 0.055 | 0.004 | 4.0E-51 | 0.718 | Yes |
| rs7832643 | 8:145094645 | PLEC1 | T/G | 0.153 | 0.148 | 16913 | -0.003 | 0.016 | 0.845 | 164854 | 0.034 | 0.004 | 7.0E-19 | 0.021 | Yes |
| rs3780181 | 9:2630759 | VLDLR | A/G | 0.910 | 0.887 | 16913 | 0.049 | 0.020 | 0.017 | 171976 | 0.045 | 0.007 | 1.0E-09 | 0.850 | Yes |
| rs1883025 | 9:106704122 | ABCA1 | C/T | 0.783 | 0.757 | 16913 | 0.017 | 0.013 | 0.207 | 172330 | 0.030 | 0.004 | 1.0E-11 | 0.363 | Yes |
| rs579459 | 9:135143989 | *ABO/SURF6 | C/T | 0.216 | 0.190 | 16913 | 0.084 | 0.013 | 1.7E-10 | 172706 | 0.067 | 0.005 | 3.0E-49 | 0.215 | Yes |
| rs2255141 | 10:113923876 | GPAM/RP11-426E5.2 | A/G | 0.312 | 0.242 | 16913 | 0.051 | 0.012 | 9.3E-06 | 173003 | 0.030 | 0.004 | 7.0E-14 | 0.080 | Yes |
| rs174532 | 11:61305450 | C11orf9 | A/G | 0.001 | 0.001 | 12601 | -0.017 | 0.123 | 0.890 | 172981 | 0.035 | 0.004 | 5.0E-17 | 0.671 | Yes |
| rs1535 | 11:61354548 | FADS2 | A/G | 0.563 | 0.434 | 16913 | 0.045 | 0.011 | 9.2E-05 | 168360 | 0.053 | 0.004 | 3.0E-43 | 0.501 | Yes |
| rs11220462 | 11:125749162 | ST3GAL4 | A/G | 0.347 | 0.377 | 16913 | 0.029 | 0.011 | 0.011 | 145030 | 0.059 | 0.006 | 3.0E-23 | 0.018 | Yes |
| rs653178 | 12:110492139 | ATXN2 | T/C | 0.997 | 0.997 | 14981 | 0.103 | 0.080 | 0.195 | 165312 | 0.023 | 0.004 | 2.0E-09 | 0.312 | Yes |
| rs6489818 | 12:110794963 | MAPKAPK5 | A/G | 0.882 | 0.847 | 16913 | -0.009 | 0.016 | 0.583 | 173044 | 0.028 | 0.005 | 6.0E-09 | 0.031 | Yes |
| rs1169288 | 12:119901033 | HNF1A | C/A | 0.396 | 0.389 | 16913 | 0.017 | 0.011 | 0.142 | 163086 | 0.038 | 0.004 | 9.0E-21 | 0.088 | Yes |
| rs4942486 | 13:31851388 | BRCA2 | T/C | 0.462 | 0.467 | 16913 | 0.037 | 0.011 | 7.9E-04 | 171930 | 0.024 | 0.004 | 3.0E-11 | 0.283 | Yes |
| rs8017377 | 14:23953727 | KIAA1305 | A/G | 0.055 | 0.050 | 16913 | 0.055 | 0.024 | 0.024 | 172866 | 0.030 | 0.004 | 3.0E-15 | 0.317 | Yes |
| rs9989419 | 16:55542640 | *CETP/HERPUD1/NLRC5/SLC12A3 | A/G | 0.236 | 0.244 | 16913 | -0.015 | 0.013 | 0.253 | 163625 | 0.028 | 0.004 | 8.0E-13 | 0.002 | Yes |
| rs2000999 | 16:70665594 | DLP/HPR | A/G | 0.265 | 0.310 | 16913 | 0.041 | 0.013 | 9.9E-04 | 171510 | 0.065 | 0.005 | 1.0E-45 | 0.078 | Yes |
| rs314253 | 17:7032374 | *ACADVL/ASGR1/CS266489/DLG4/DVL2/PHF23 | T/C | 0.516 | 0.548 | 16913 | 0.029 | 0.011 | 0.008 | 169706 | 0.024 | 0.004 | 2.0E-10 | 0.672 | Yes |
| rs4791641 | 17:8101874 | PFAS | C/T | 0.749 | 0.773 | 16913 | -0.011 | 0.013 | 0.377 | 171304 | 0.020 | 0.004 | 4.0E-08 | 0.017 | Yes |
| rs7225700 | 17:42746803 | *AX748120/C17orf57/ITGB3 | C/T | 0.669 | 0.651 | 16913 | 0.014 | 0.012 | 0.247 | 171505 | 0.030 | 0.004 | 8.0E-15 | 0.187 | Yes |
| rs6511720 | 19:11063306 | LDLR | G/T | 0.993 | 0.988 | 16913 | 0.192 | 0.064 | 0.002 | 170608 | 0.221 | 0.006 | 3.0E-289 | 0.651 | Yes |

| | | | | | | CKB Univariate Model | | | | GLGC (As Used for Score) | | | | | |
|---|---|---|---|---|---|---|---|---|---|---|---|---|---|---|---|
| rsID | Chr:Pos | Genes | Effect/ other allele | EAF (CKB) | EAF (1KG) | N | Estimate (SD) | SE | P-value | N | Estimate (SD) | SE | P-value | Het P-value | Included in CKB LDL-C GS? |
| rs688 | 19:11088602 | LDLR | T/C | 0.167 | 0.184 | 16913 | 0.077 | 0.014 | 1.1E-07 | 166792 | 0.054 | 0.004 | 9.0E-48 | 0.130 | Yes |
| rs7254892 | 19:50081436 | PVRL2 | G/A | 0.934 | 0.916 | 16913 | 0.703 | 0.021 | 5.7E-236 | 139198 | 0.485 | 0.012 | 3.8E-357 | 2.0E-19* | Yes |
| rs492602 | 19:53898229 | FUT2 | G/A | 0.007 | 0.004 | 16913 | 0.047 | 0.059 | 0.427 | 170009 | 0.029 | 0.004 | 3.0E-14 | 0.763 | Yes |
| rs364585 | 20:12910718 | *SPTLC3 | G/A | 0.579 | 0.567 | 16913 | -0.009 | 0.011 | 0.390 | 171526 | 0.025 | 0.004 | 4.0E-11 | 0.003 | Yes |
| rs2328223 | 20:17793921 | None | C/A | 0.208 | 0.225 | 16913 | 0.020 | 0.013 | 0.140 | 170762 | 0.030 | 0.005 | 2.0E-09 | 0.484 | Yes |
| rs7264396 | 20:33618155 | FER1L4 | C/T | 0.776 | 0.782 | 16913 | 0.008 | 0.013 | 0.522 | 171527 | 0.025 | 0.005 | 3.0E-08 | 0.241 | Yes |
| rs6016381 | 20:38613850 | None | T/C | 0.424 | 0.407 | 16913 | 0.019 | 0.011 | 0.090 | 171556 | 0.036 | 0.004 | 6.0E-22 | 0.147 | Yes |
| rs6065311 | 20:39157752 | TOP1 | C/T | 0.820 | 0.806 | 16913 | 0.018 | 0.014 | 0.209 | 171333 | 0.042 | 0.004 | 3.0E-30 | 0.102 | Yes |
| rs1800961 | 20:42475778 | HNF4A | C/T | 0.985 | 0.984 | 16913 | 0.027 | 0.044 | 0.541 | 142698 | 0.069 | 0.011 | 1.0E-10 | 0.362 | Yes |
| rs5763662 | 22:28708703 | MTMR3 | T/C | 0.096 | 0.115 | 16913 | 0.025 | 0.019 | 0.189 | 162777 | 0.077 | 0.012 | 2.0E-10 | 0.020 | Yes |
| rs1010167 | 1:110000250 | GSTM2/GSTM4 | G/C | 0.654 | 0.639 | 16913 | -0.019 | 0.012 | 0.109 | 165771 | 0.025 | 0.004 | 3.0E-10 | 3.7E-04* | No |
| rs515135 | 2:21139562 | *APOB | C/T | 0.897 | 0.917 | 16913 | -0.009 | 0.018 | 0.604 | 173029 | 0.139 | 0.005 | 1.0E-188 | 8.9E-16* | No |
| rs7640978 | 3:32508014 | CMTM6 | C/T | 0.942 | 0.930 | 16913 | -0.047 | 0.023 | 0.044 | 172228 | 0.039 | 0.007 | 1.0E-08 | 3.9E-04* | No |
| rs4530754 | 5:122883315 | CSNK1G3 | A/G | 0.352 | 0.343 | 16913 | -0.015 | 0.011 | 0.187 | 173003 | 0.028 | 0.004 | 4.0E-14 | 3.5E-04* | No |
| rs2297374 | 6:160495975 | SLC22A1 | C/T | 0.658 | 0.664 | 16913 | -0.013 | 0.012 | 0.263 | 172978 | 0.033 | 0.004 | 6.0E-18 | 1.8E-04* | No |
| rs2073547 | 7:44548856 | *DDX21/DDX56/DQ574505/NPC1L1/TMED4 | G/A | 0.369 | 0.375 | 16913 | -0.003 | 0.011 | 0.780 | 169889 | 0.049 | 0.005 | 5.0E-23 | 2.6E-05* | No |
| rs10832962 | 11:18612847 | *AK097654/AL137366/SPTY2D1/UEVLD | T/C | 0.489 | 0.491 | 16913 | -0.010 | 0.011 | 0.363 | 172920 | 0.032 | 0.004 | 2.0E-15 | 3.0E-04* | No |
| rs10790162 | 11:116144314 | BUD13 | A/G | 0.212 | 0.235 | 16913 | -0.034 | 0.013 | 0.011 | 173007 | 0.076 | 0.007 | 3.0E-26 | 3.7E-13* | No |
| rs10401969 | 19:19268718 | SF4 | T/C | 0.906 | 0.893 | 16913 | -0.034 | 0.019 | 0.075 | 171477 | 0.118 | 0.007 | 2.0E-60 | 7.4E-14* | No |
| rs6859 | 19:50073874 | PVRL2 | A/G | 0.315 | 0.309 | 16913 | -0.040 | 0.012 | 6.5E-04 | 166756 | 0.084 | 0.004 | 1.0E-101 | 1.1E-22* | No |

CKB estimates are for rank inverse normal transformed LDL-C, after adjustment for age, age$^2$, fasting time, fasting time$^2$, ascertainment group, sex, and 1-9 regional principal components, stratified by region.
Exclusion criteria: Bonferroni corrected heterogeneity p-value < 0.05/74 and either non-significant CKB association (p-value > 0.05/74) OR non-concordant direction of effect for the CKB lipid effect estimate
* in genes column indicates genes were found within 50KB, used where no genes overlap SNP location
* in Het P-value column indicates significant heterogeneity after Bonferroni correction

**Supplementary Table 3:** ICD-10 codes of vascular and non-vascular endpoints in CKB

**Vascular outcomes**

- Major coronary events (MCE): Fatal IHD (ICD-10: I20-I25) or non-fatal myocardial infarction (ICD-10: I21) or coronary revascularisation procedures.
- Fatal or non-fatal myocardial infarction (ICD-10: I21).
- Fatal or non-fatal ischaemic stroke (ICD-10: I63).
- Fatal or non-fatal intracerebral haemorrhage (ICD-10: 161).
- Fatal or non-fatal stroke (ICD-10: I60, I61, 163, I64).
- Occlusive CVD: Fatal IHD (ICD-10: I20-I25) or non-fatal myocardial infarction (ICD-10: I21) or coronary revascularisation procedures or fatal or non-fatal ischaemic stroke (ICD-10: I63).
- Fatal cardiovascular disease (ICD-10: I00–99).
- Major vascular events (MVE): Fatal or non-fatal myocardial infarction (ICD-10: I21) or coronary revascularisation procedures or fatal or non-fatal stroke (ICD-10: I60, I61, 163, I64) or fatal cardiovascular disease (ICD-10: I00–I99).
- **Common vascular controls**: Exclude prevalent CHD, prevalent stroke or transient ischaemic attack or incident MVE.

**Main non-vascular outcomes**

- Diabetes (ICD-10: E10 – E14), including prevalent and screen-detected diabetes.
- Chronic kidney disease (CKD) (ICD-10: E10.2+, E11.2+, E12.2+, E13.2+, E14.2+, I12.0, I12.9, I13.0, I13.1, I13.2, I13.9, M10.3, M32.1+, N02, N03, N04, N05, N08.3*, N11, N12, N13, N15, N18, N19, N25, N26, N27.1, N27.9, N28.9, O10.2, O10.3, R94.4, T86.1, Z94.0). Exclude prevalent kidney disease from controls.
- Chronic liver disease including cirrhosis and hepatitis (ICD-10: CD-10 B18, B19, B25.1+, B58.1+, B94.2, K72, K73, K74, K75, K76, K77*, Z22.5) Exclude prevalent cirrhosis or hepatitis from controls.
- Malignant neoplasms (ICD-10: C00-C97). Exclude prevalent cancer from controls.
- Eye diseases (ICD-10: H00-H59).
- COPD (ICD-10: J41-J44)
- Non-vascular mortality (all ICD-10 codes excluding I00-I99).

**Respiratory outcomes**

- Chronic obstructive pulmonary disease (COPD)
  - Incident COPD: ICD-10: J41 –J44.
  - Prevalent COPD: COPD at baseline according to spirometry measures and Global Lung Initiative definition of FEV1/FVC less than lower limit of normal.
- COPD Exacerbation: Incident events in individuals with COPD at baseline.
- Upper Respiratory Tract Infection (URTI): J00-J06, first event occuring 2009 or later.

**Supplementary Table 4:** ICD-10 codes of endpoints included in the phenome scan

| Endpoint description (full) | Endpoint description (short) | ICD-10 Codes |
|---|---|---|
| Certain infections and parasitic diseases | Infectious and parasitic diseases | A00-B99 |
| Malignant neoplasms (excluding bronchus and lung) | Malignant neoplasms (excluding bronchus and lung), | C00-C33, C37-C99 |
| Malignant neoplasms of bronchus and lung | Malignant neoplasms of bronchus and lung, | C34 |
| In situ, benign, uncertain or unknown behaviour neoplasms | In situ, benign, uncertain or unknown behaviour neoplasms | D00-D48 |
| Diseases of the blood and blood-forming organs and certain disorders involving the immune mechanism | Diseases of the blood | D50-D89 |
| Disorders of thyroid gland, Other disorders of glucose regulation and pancreatic internal secretion, Disorders of other endocrine glands | Other endocrine disorders | E00-E07, E15-E35 |
| Diabetes mellitus | Diabetes | E10-E14 |
| Malnutrition, Other nutritional deficiencies, Obesity and other hyperalimentation, Metabolic disorders | Malnutrition or other nutritional deficiencies | E40-E77, E79-E90 |
| Disorders of lipoprotein metabolism and other lipidaemias | Disorders of lipoprotein metabolism | E78 |
| Mental and behavioural disorders | Mental and behavioural disorders, | F00-F99 |
| Diseases of the nervous system (excluding TIA) | Diseases of the nervous system | G00-G44, G46-G99 |
| Transient cerebral ischaemic attacks and related syndromes | Transient cerebral ischaemic attacks | G45 |
| Diseases of the eye and adnexa | Diseases of the eye and adnexa | H00-H59 |
| Diseases of the ear and mastoid process | Diseases of the ear and mastoid process | H60-H95 |
| Diseases of the circulatory system (excluding hypertension, IHD, cerebrovascular disease) | Diseases of the circulatory system | I00-I09, I11-I15, I26-I52, I70-I99 |
| Essential (primary) hypertension | Essential hypertension | I10 |
| Ischaemic heart diseases | Ischaemic heart disease | I120-I25 |
| Cerebrovascular diseases | Cerebrovascular disease | I60-I69 |
| Acute upper respiratory infections | Acute Upper Respiratory Infections | J00-J06 |
| Influenza and pneumonia | Influenza and pneumonia | J09-J18 |
| Other acute lower respiratory infections | Other acute lower respiratory infections | J20-J22 |
| Other diseases of upper respiratory tract | Other diseases of upper respiratory tract | J30-J39 |

| Endpoint description (full) | Endpoint description (short) | ICD-10 Codes |
|---|---|---|
| Chronic lower respiratory diseases (excluding asthma) | Chronic lower respiratory disease | J40-J44, J47 |
| Asthma, status asthmaticus | Asthma | J45-J46 |
| Other diseases of the respiratory system | Other diseases of the respiratory system, | J60-J99 |
| Diseases of oral cavity, salivary glands and jaws (excluding gingivitis and periodontal diseases) | Diseases of oral cavity | K00-K04, K06-K14 |
| Gingivitis and periodontal diseases | Gingivitis and periodontal diseases | K05 |
| Diseases of oesophagus, stomach and duodenum | Diseases of oesophagus, stomach and duodenum | K20-K31 |
| Diseases of appendix, Hernia, Other diseases of intestines, Diseases of peritoneum, Other diseases of the digestive system | Other digestive system diseases | K35-K46, K55-K67, K90-K93 |
| Crohn disease [regional enteritis], Ulcerative colitis, Other noninfective gastroenteritis and colitis | Noninfective enteritis and colitis | K50-K52 |
| Diseases of liver | Diseases of the liver | K70-K77 |
| Disorders of gallbladder, biliary tract and pancreas | Disorders of biliary tract and pancreas | K80-K87 |
| Diseases of the skin and subcutaneous tissue | Diseases of the skin | L00-L99 |
| Arthropathies | Arthropathies | M00-M25 |
| Diseases of the musculoskeletal system and connective tissue (excluding arthropothies) | Other musculoskeletal disorders | M30-M99 |
| Glomerular diseases, Renal tubulo-interstitial diseases, Renal failure, Urolithiasis, Other disorders of kidney and ureter | Renal disorders | N00-N29 |
| Other diseases of urinary system | Other diseases of urinary system | N30-N39 |
| Diseases of male genital organs | Diseases of the male genital organs | N40-N51 |
| Disorders of breast | Disorders of breast | N60-N64 |
| Inflammatory diseases of female pelvic organs | Inflammatory diseases of female pelvic organs | N70-N77 |
| Non-inflammatory disorders of female genital tract, Other disorders of the genitourinary system | Other female genitourinary diseases | N80-N99 |

**Supplementary Table 5:** Numbers and ascertainments of cases and controls included in analyses of disease associations with the *PCSK9* genetic score

| | | Ascertainment Group | | | | | | | | |
|---|---|---|---|---|---|---|---|---|---|---|
| | | **Population Representative** | **IS case** | **MI case** | **Fatal IHD case** | **ICH case** | **SAH case** | **COPD** | **Other** | **Total** |
| **Group** | **Endpoint** | **Cases/ Controls** | **Cases/ Controls** | **Cases/ Controls** | **Cases/ Controls** | **Cases/ Controls** | **Cases/ Controls** | **Cases/ Controls** | **Cases/ Controls** | **Cases/ Controls** |
| Figure 2: Cardiovascular diseases | Major Coronary Events | 2390/59421 | X/X | 1657/X | 696/X | X/X | X/X | X/X | 277/X | 5020/59421 |
| | Ischaemic Stroke | 6938/59421 | 4064/X | X/X | X/X | X/X | X/X | X/X | 465/X | 11467/59421 |
| | Intracerebral haemorrhage | 1102/59421 | X/X | X/X | X/X | 4680/X | X/X | X/X | 124/X | 5906/59421 |
| | Major Occlusive Vascular Events | 8631/59421 | 4064/X | 1657/X | 696/X | X/X | X/X | X/X | 704/X | 15752/59421 |
| | Fatal Cardiovascular Disease | 2161/59421 | 118/X | 961/X | 696/X | 2340/X | 44/X | X/X | 323/X | 6643/59421 |
| | Major Vascular Events | 9643/59421 | 4064/X | 1657/X | 696/X | 4680/X | 372/X | X/X | 867/X | 21979/59421 |
| Figure 2: Other diseases | Diabetes | 6863/64051 | X/X | X/X | X/X | X/X | X/X | X/X | 605/X | 7468/64051 |
| | Chronic Kidney Disease | 963/68918 | X/X | X/X | X/X | X/X | X/X | X/X | 117/X | 1080/68918 |
| | Chronic Liver Disease | 607/69492 | X/X | X/X | X/X | X/X | X/X | X/X | 72/X | 679/69492 |
| | Malignant Neoplasms | 4099/66593 | X/X | X/X | X/X | X/X | X/X | X/X | 462/X | 4561/66593 |
| | Diseases of Eye and Adnexa | 3114/67800 | X/X | X/X | X/X | X/X | X/X | X/X | 440/X | 3554/67800 |
| | Incident COPD | 2559/66997 | X/X | X/X | X/X | X/X | X/X | 3727/X | 550/X | 6836/66997 |
| | Non-Vascular Mortality | 3758/67156 | X/X | X/X | X/X | X/X | X/X | X/X | 627/X | 4385/67156 |
| Figure 3: UKB repliication | Acute upper respiratory infections | 980/69934 | X/X | X/X | X/X | X/X | X/X | X/X | 115/X | 1095/69934 |
| | Asthma diagnosed by doctor | 389/70525 | X/X | X/X | X/X | X/X | X/X | X/X | 38/X | 427/70525 |
| | Mood (affective) disorders | 132/70782 | X/X | X/X | X/X | X/X | X/X | X/X | 5/X | 137/70782 |
| | Dorsopathies not classified as deforming | 2198/68716 | X/X | X/X | X/X | X/X | X/X | X/X | 244/X | 2442/68716 |
| | Cerebrovascular diseases | 9681/61233 | X/X | X/X | X/X | X/X | X/X | X/X | X/X | 9681/61233 |
| | Soft tissue disorders including bursitis | 291/70623 | X/X | X/X | X/X | X/X | X/X | X/X | 39/X | 330/70623 |
| | Malignant neoplasm of breast | 389/41954 | X/X | X/X | X/X | X/X | X/X | X/X | 25/X | 414/41954 |

| | | Ascertainment Group | | | | | | | | |
|---|---|---|---|---|---|---|---|---|---|---|
| | | Population Representative | IS case | MI case | Fatal IHD case | ICH case | SAH case | COPD | Other | Total |
| Group | Endpoint | Cases/ Controls | Cases/ Controls | Cases/ Controls | Cases/ Controls | Cases/ Controls | Cases/ Controls | Cases/ Controls | Cases/ Controls | Cases/ Controls |
| Figure 5: Respiratory Disease | Prevalent COPD | 4539/52355 | X/X | X/X | X/X | X/X | X/X | X/X | 566/X | 5105/52355 |
| | Incident COPD | 2559/66997 | X/X | X/X | X/X | X/X | X/X | 3727/X | 550/X | 6836/66997 |
| | Incident COPD (Fatal) | 511/66997 | X/X | X/X | X/X | X/X | X/X | 971/X | 150/X | 1632/66997 |
| | Incident COPD (Non-Fatal) | 2048/66997 | X/X | X/X | X/X | X/X | X/X | 2756/X | 400/X | 5204/66997 |
| | COPD Exacerbations | 754/3785 | X/X | X/X | X/X | X/X | X/X | 1304/X | 154/412 | 2212/4197 |
| | COPD Exacerbations (Fatal) | 208/3785 | X/X | X/X | X/X | X/X | X/X | 476/X | 46/412 | 730/4197 |
| | COPD Exacerbations (Non-Fatal) | 546/3785 | X/X | X/X | X/X | X/X | X/X | 828/X | 108/412 | 1482/4197 |
| | Acute upper respiratory infections | 980/69934 | X/X | X/X | X/X | X/X | X/X | X/X | 115/X | 1095/69934 |
| Figure S7: Phenome-wide Scan | Infectious and parasitic diseases | 2232/68682 | X/X | X/X | X/X | X/X | X/X | X/X | 343/X | 2575/68682 |
| | Malignant neoplasms (excluding bronchus and lung), | 3492/67422 | X/X | X/X | X/X | X/X | X/X | X/X | 365/X | 3857/67422 |
| | Malignant neoplasms of bronchus and lung, | 884/70030 | X/X | X/X | X/X | X/X | X/X | X/X | 125/X | 1009/70030 |
| | In situ, benign, uncertain or unknown behaviour neoplasms, | 2034/68880 | X/X | X/X | X/X | X/X | X/X | X/X | 144/X | 2178/68880 |
| | Diseases of the blood | 595/70319 | X/X | X/X | X/X | X/X | X/X | X/X | 68/X | 663/70319 |
| | Other endocrine disorders | 645/70269 | X/X | X/X | X/X | X/X | X/X | X/X | 42/X | 687/70269 |
| | Diabetes mellitus | 4971/65943 | X/X | X/X | X/X | X/X | X/X | X/X | 424/X | 5395/65943 |
| | Malnutrition or other nutritional deficiencies | 446/70468 | X/X | X/X | X/X | X/X | X/X | X/X | 67/X | 513/70468 |
| | Disorders of lipoprotein metabolism | 535/70379 | X/X | X/X | X/X | X/X | X/X | X/X | 45/X | 580/70379 |
| | Mental and behavioural disorders, | 730/70184 | X/X | X/X | X/X | X/X | X/X | X/X | 70/X | 800/70184 |
| | Diseases of the nervous system | 1347/69567 | X/X | X/X | X/X | X/X | X/X | X/X | 134/X | 1481/69567 |
| | Transient cerebral ischaemic attacks | 2992/67922 | X/X | X/X | X/X | X/X | X/X | X/X | 270/X | 3262/67922 |
| | Diseases of the eye and adnexa | 3114/67800 | X/X | X/X | X/X | X/X | X/X | X/X | 440/X | 3554/67800 |

| | | Ascertainment Group | | | | | | | | |
|---|---|---|---|---|---|---|---|---|---|---|
| | | Population Representative | IS case | MI case | Fatal IHD case | ICH case | SAH case | COPD | Other | Total |
| Group | Endpoint | Cases/ Controls | Cases/ Controls | Cases/ Controls | Cases/ Controls | Cases/ Controls | Cases/ Controls | Cases/ Controls | Cases/ Controls | Cases/ Controls |
| | Diseases of the ear and mastoid process | 697/70217 | X/X | X/X | X/X | X/X | X/X | X/X | 61/X | 758/70217 |
| | Diseases of the circulatory system | 5082/65832 | X/X | X/X | X/X | X/X | X/X | X/X | 634/X | 5716/65832 |
| | Essential hypertension | 5500/65414 | X/X | X/X | X/X | X/X | X/X | X/X | 673/X | 6173/65414 |
| | Ischaemic heart disease | 7548/63366 | X/X | X/X | X/X | X/X | X/X | X/X | 779/X | 8327/63366 |
| | Cerebrovascular disease | 9681/61233 | X/X | X/X | X/X | X/X | X/X | X/X | 869/X | 10550/61233 |
| | Acute upper respiratory infections | 1126/69788 | X/X | X/X | X/X | X/X | X/X | X/X | 125/X | 1251/69788 |
| | Influenza and pneumonia | 3901/67013 | X/X | X/X | X/X | X/X | X/X | X/X | 534/X | 4435/67013 |
| | Other acute lower respiratory infections | 1089/69825 | X/X | X/X | X/X | X/X | X/X | X/X | 150/X | 1239/69825 |
| | Other diseases of upper respiratory tract | 635/70279 | X/X | X/X | X/X | X/X | X/X | X/X | 44/X | 679/70279 |
| | Chronic lower respiratory disease | 3271/67643 | X/X | X/X | X/X | X/X | X/X | X/X | 617/X | 3888/67643 |
| | Asthma | 353/70561 | X/X | X/X | X/X | X/X | X/X | X/X | 50/X | 403/70561 |
| | Other diseases of the respiratory system, | 777/70137 | X/X | X/X | X/X | X/X | X/X | X/X | 113/X | 890/70137 |
| | Diseases of oral cavity | 147/70767 | X/X | X/X | X/X | X/X | X/X | X/X | 8/X | 155/70767 |
| | Gingivitis and periodontal diseases | 34/70880 | X/X | X/X | X/X | X/X | X/X | X/X | 6/X | 40/70880 |
| | Diseases of oesophagus, stomach and duodenum | 2995/67919 | X/X | X/X | X/X | X/X | X/X | X/X | 397/X | 3392/67919 |
| | Other digestive system diseases | 3032/67882 | X/X | X/X | X/X | X/X | X/X | X/X | 288/X | 3320/67882 |
| | Noninfective enteritis and colitis | 220/70694 | X/X | X/X | X/X | X/X | X/X | X/X | 38/X | 258/70694 |
| | Diseases of the liver | 829/70085 | X/X | X/X | X/X | X/X | X/X | X/X | 85/X | 914/70085 |
| | Disorders of biliary tract and pancreas | 2470/68444 | X/X | X/X | X/X | X/X | X/X | X/X | 298/X | 2768/68444 |
| | Diseases of the skin | 649/70265 | X/X | X/X | X/X | X/X | X/X | X/X | 86/X | 735/70265 |
| | Arthropathies | 1461/69453 | X/X | X/X | X/X | X/X | X/X | X/X | 172/X | 1633/69453 |
| | Other musculoskeletal disorders | 4689/66225 | X/X | X/X | X/X | X/X | X/X | X/X | 557/X | 5246/66225 |

| | | Ascertainment Group | | | | | | | | |
|---|---|---|---|---|---|---|---|---|---|---|
| | | Population Representative | IS case | MI case | Fatal IHD case | ICH case | SAH case | COPD | Other | Total |
| Group | Endpoint | Cases/ Controls | Cases/ Controls | Cases/ Controls | Cases/ Controls | Cases/ Controls | Cases/ Controls | Cases/ Controls | Cases/ Controls | Cases/ Controls |
| | Renal disorders | 2621/68293 | X/X | X/X | X/X | X/X | X/X | X/X | 335/X | 2956/68293 |
| | Other diseases of urinary system | 736/70178 | X/X | X/X | X/X | X/X | X/X | X/X | 92/X | 828/70178 |
| | Diseases of the male genital organs | 653/27918 | X/X | X/X | X/X | X/X | X/X | X/X | 136/X | 789/27918 |
| | Disorders of breast (females only) | 211/42132 | X/X | X/X | X/X | X/X | X/X | X/X | 10/X | 221/42132 |
| | Inflammatory diseases of female pelvic organs | 573/41770 | X/X | X/X | X/X | X/X | X/X | X/X | 56/X | 629/41770 |
| | Other female genitourinary diseases | 884/41459 | X/X | X/X | X/X | X/X | X/X | X/X | 55/X | 939/41459 |

**Supplementary Table 6:** Disease definitions for replication of outcomes reported as associated with a functional *PCSK9* variant in UK Biobank

| **Outcome description** | **Case definition** |
|---|---|
| Acute upper respiratory infections | ICD-10 J00-J06 |
| Asthma diagnosed by doctor | self-report |
| Mood (affective) disorders | ICD-10 F30-F39 |
| Dorsopathies not classified as deforming | ICD-10 M50-M54 |
| Cerebrovascular diseases | ICD-10 I60-I69 |
| Soft tissue disorders including bursitis | ICD-10 M70-M79 |
| Malignant neoplasm of breast | ICD-10 C50, females only |

**Supplementary Table 7:** Association of the *PCSK9* genetic score with carotid intima media thickness and presence of carotid plaque in CKB second resurvey participants

| Phenotype | N | Effect (SD)* | SE | OR* | 95% CI | | P |
|---|---|---|---|---|---|---|---|
| | | | | | LL | UL | |
| Mean CIMT | 20896 | -0.24 | 0.06 | | –0.36 | –0.12 | 0.00012 |
| Plaque | 8340 cases<br>12555 controls | | | 0.61 | 0.45 | 0.83 | 0.0015 |

*Effect/OR expressed per 1 SD lower LDL-C

**Supplementary Table 8:** Association of the *PCSK9* genetic score with FEV1/FVC measured at baseline in CKB participants

| SNP | Effect/ other allele | Effect allele freq. | N | effect (SD)* | SE | P-value |
|---|---|---|---|---|---|---|
| rs151193009 | C/T | 0.987 | 83715 | -0.00483 | 0.00364 | 0.185 |
| rs2495477 | A/G | 0.738 | 83715 | 0.01503 | 0.01088 | 0.167 |
| rs11206517 | G/T | 0.060 | 83715 | -0.00448 | 0.01096 | 0.683 |
| Meta-analysis | - | - | - | -0.00298 | 0.00329 | 0.366 |

*Effect expressed per 1 SD lower LDL-C

**Supplementary Table 9:** Association of a loss-of-function *PCSK9* SNP with exacerbations of COPD in UK Biobank

| SNP | Chr:bp | Effect/other allele | EAF | Per-allele log odds (SE) of COPD exacerbation | OR of COPD exacerbation per 1-SD lower LDL-C |
|---|---|---|---|---|---|
| rs11591147 | 1:55505647 | G/T | 0.983 | 0.005 (0.005) | 0.55 (0.20, 1.50) |

### Supplementary Figures

**Supplementary Figure 1.** Violin plots showing the distribution of measured FEV1/FVC in each CKB region

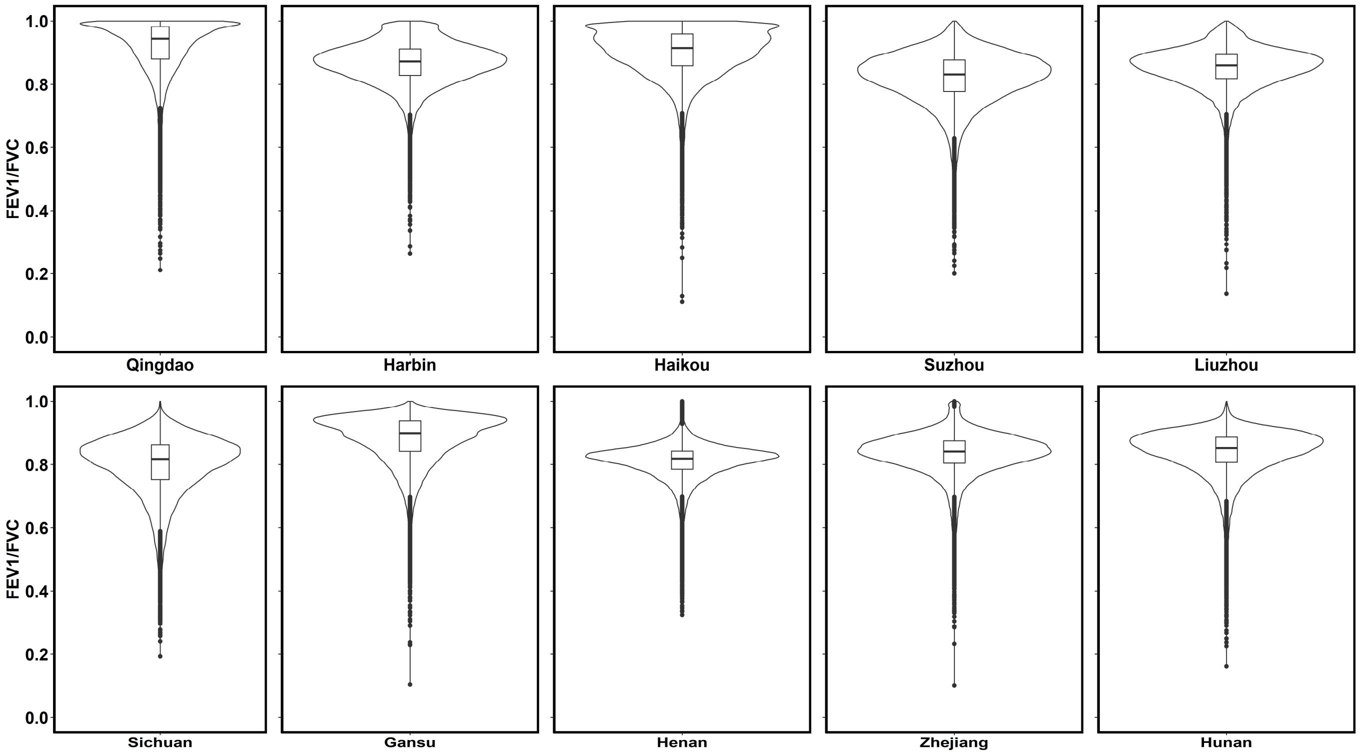

**Supplementary Figure 2.** Histogram of measured FEV1/FVC by prevalent COPD status.

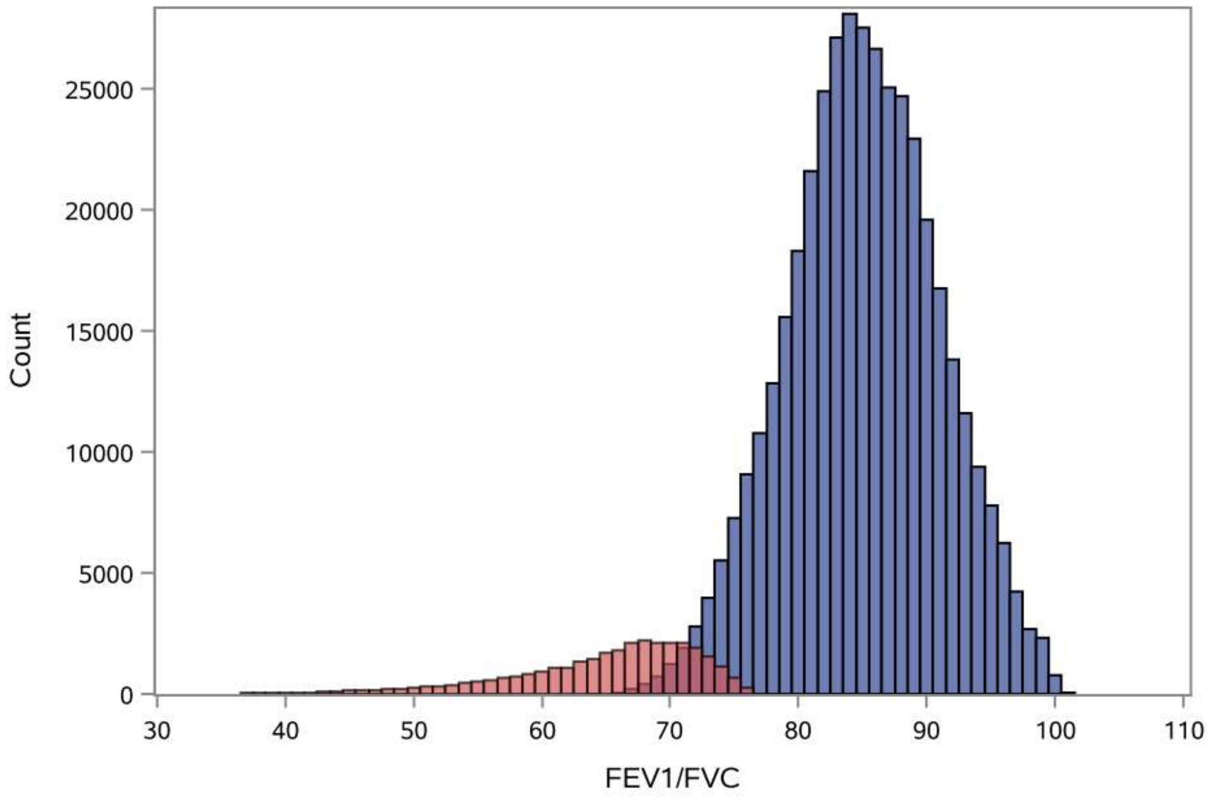

COPD is defined as FEV1/FVC below the lower limit of normal predicted from Global Lung Initiative reference equations[37], derived using ancestry, age, height, and sex.

**Supplementary Figure 3:** Sub-group analyses of association of PCSK9 genetic score with LDL-cholesterol.

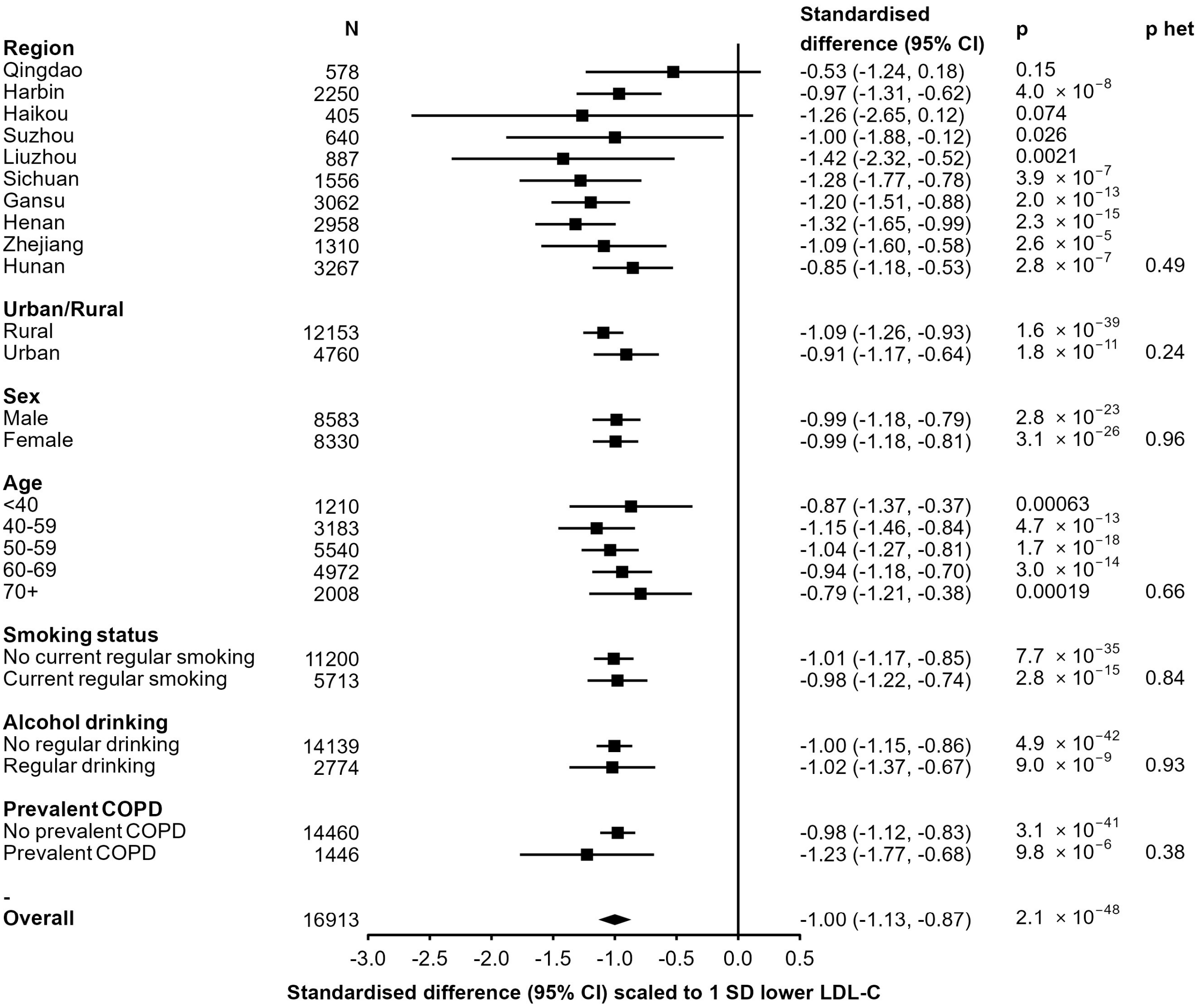

Analysis of prevalent COPD limited to 8 geographical regions in which spirometry passed quality control.

**Supplementary Figure 4:** Association of *PCSK9* genetic score with metabolic biomarkers quantified by [1]H-NMR spectroscopy.

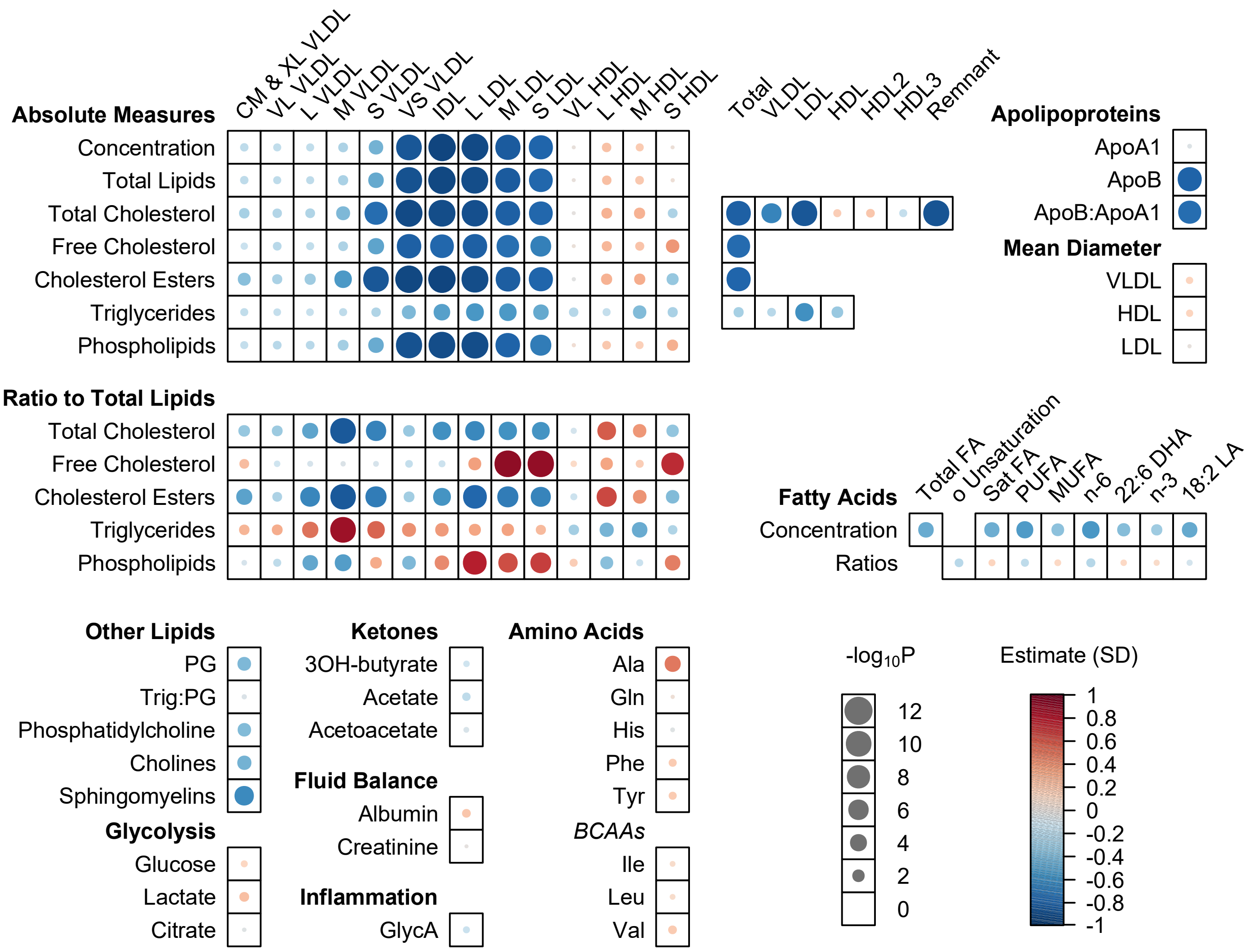

Heatmap representation of metabolome associations with the *PCSK9* genetic score was generated using the R package *bubblePlot*.

**Supplementary Figure 5:** Association of *PCSK9* genetic score with non-lipid blood measures

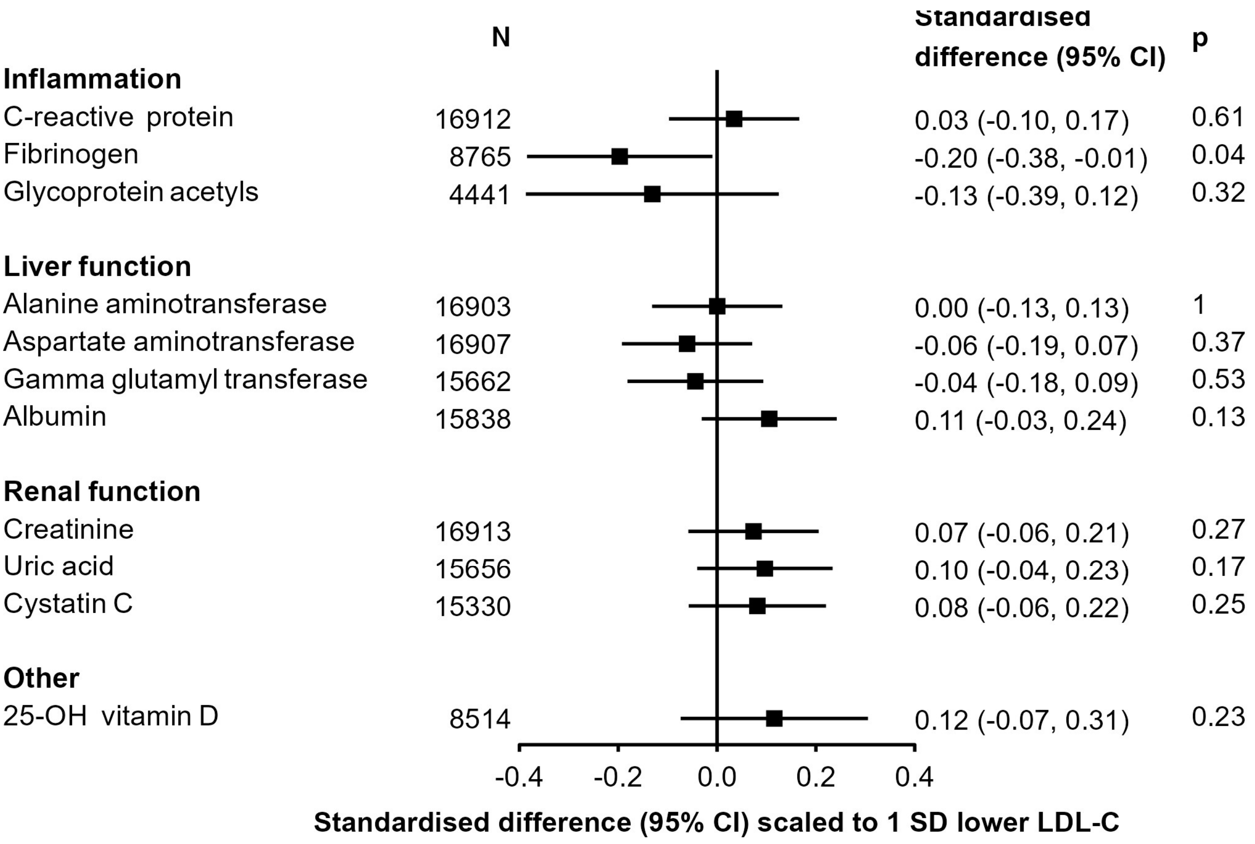

Glycoprotein acetyl measurements were derived from $^{1}$H-NMR metabolomics, other measures were from clinical biochemistry assays.

**Supplementary Figure 6:** Phenome-wide scan of *PCSK9* genetic score associations in China Kadoorie Biobank

| | Cases/Controls | OR (95% CI) | p |
|---|---|---|---|
| Infectious and parasitic diseases | 2575/68682 | 1.28 (0.91, 1.81) | 0.16 |
| Malignant neoplasms (excluding bronchus and lung) | 3857/67422 | 1.04 (0.78, 1.39) | 0.77 |
| Malignant neoplasms of bronchus and lung | 1009/70030 | 1.09 (0.64, 1.87) | 0.75 |
| In situ, benign, uncertain or unknown behaviour neoplasms | 2178/68880 | 0.91 (0.62, 1.35) | 0.65 |
| Diseases of the blood | 663/70319 | 1.66 (0.88, 3.16) | 0.12 |
| Other endocrine disorders | 687/70269 | 1.34 (0.71, 2.51) | 0.37 |
| Diabetes mellitus | 5395/65943 | 0.94 (0.73, 1.21) | 0.62 |
| Malnutrition or other nutritional deficiencies | 513/70468 | 1.06 (0.46, 2.42) | 0.9 |
| Disorders of lipoprotein metabolism | 580/70379 | 0.69 (0.32, 1.51) | 0.36 |
| Mental and behavioural disorders | 800/70184 | 1.04 (0.57, 1.93) | 0.89 |
| Diseases of the nervous system | 1481/69567 | 1.37 (0.89, 2.10) | 0.15 |
| Transient cerebral ischaemic attacks | 3262/67922 | 0.95 (0.70, 1.30) | 0.76 |
| Diseases of the eye and adnexa | 3554/67800 | 0.98 (0.72, 1.33) | 0.87 |
| Diseases of the ear and mastoid process | 758/70217 | 0.91 (0.47, 1.77) | 0.78 |
| Diseases of the circulatory system | 5716/65832 | 1.11 (0.87, 1.41) | 0.41 |
| Essential hypertension | 6173/65414 | 0.89 (0.70, 1.13) | 0.34 |
| Ischaemic heart disease | 8327/63366 | 1.00 (0.81, 1.23) | 0.99 |
| Cerebrovascular disease | 10550/61233 | 0.90 (0.74, 1.10) | 0.3 |
| Acute upper respiratory infections | 1251/69788 | 1.98 (1.25, 3.13) | 0.0034 |
| Influenza and pneumonia | 4435/67013 | 0.93 (0.70, 1.23) | 0.61 |
| Other acute lower respiratory infections | 1239/69825 | 1.00 (0.59, 1.68) | 0.99 |
| Other diseases of upper respiratory tract | 679/70279 | 1.17 (0.60, 2.28) | 0.64 |
| Chronic lower respiratory disease | 3888/67643 | 1.08 (0.80, 1.48) | 0.61 |
| Asthma | 403/70561 | 0.59 (0.22, 1.56) | 0.29 |
| Other diseases of the respiratory system | 890/70137 | 1.06 (0.58, 1.92) | 0.85 |
| Diseases of oral cavity | 155/70767 | 1.11 (0.24, 5.12) | 0.89 |
| Gingivitis and periodontal diseases | 40/70880 | 0.21 (0.00, 9.74) | 0.42 |
| Diseases of oesophagus, stomach and duodenum | 3392/67919 | 1.12 (0.82, 1.54) | 0.47 |
| Other digestive system diseases | 3320/67882 | 1.12 (0.83, 1.53) | 0.46 |
| Noninfective enteritis and colitis | 258/70694 | 3.25 (1.36, 7.74) | 0.0079 |
| Diseases of the liver | 914/70085 | 1.10 (0.62, 1.96) | 0.74 |
| Disorders of biliary tract and pancreas | 2768/68444 | 0.90 (0.63, 1.28) | 0.57 |
| Diseases of the skin | 735/70265 | 1.02 (0.53, 1.94) | 0.96 |
| Arthropathies | 1633/69453 | 1.16 (0.75, 1.81) | 0.5 |
| Other musculoskeletal disorders | 5246/66225 | 1.13 (0.87, 1.45) | 0.36 |
| Renal disorders | 2956/68293 | 0.73 (0.51, 1.04) | 0.084 |
| Other diseases of urinary system | 828/70178 | 1.25 (0.70, 2.23) | 0.45 |
| Diseases of the male genital organs | 789/27918 | 1.58 (0.86, 2.92) | 0.14 |
| Disorders of breast (female) | 221/42132 | 3.34 (1.21, 9.22) | 0.02 |
| Inflammatory diseases of female pelvic organs | 629/41770 | 1.03 (0.49, 2.16) | 0.94 |
| Other female genitourinary diseases | 939/41459 | 1.08 (0.60, 1.95) | 0.79 |

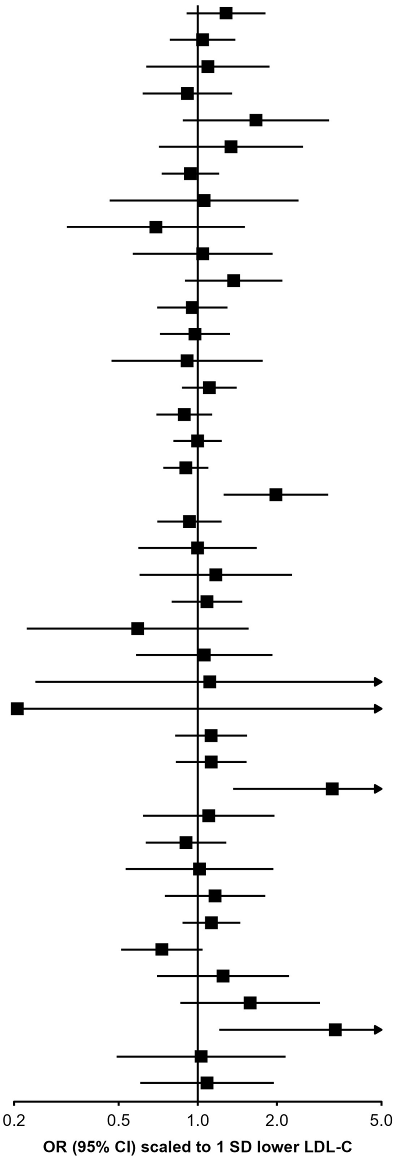

**Supplementary Figure 7:** Sub-group analyses of association of *PCSK9* genetic score with upper respiratory tract infections.

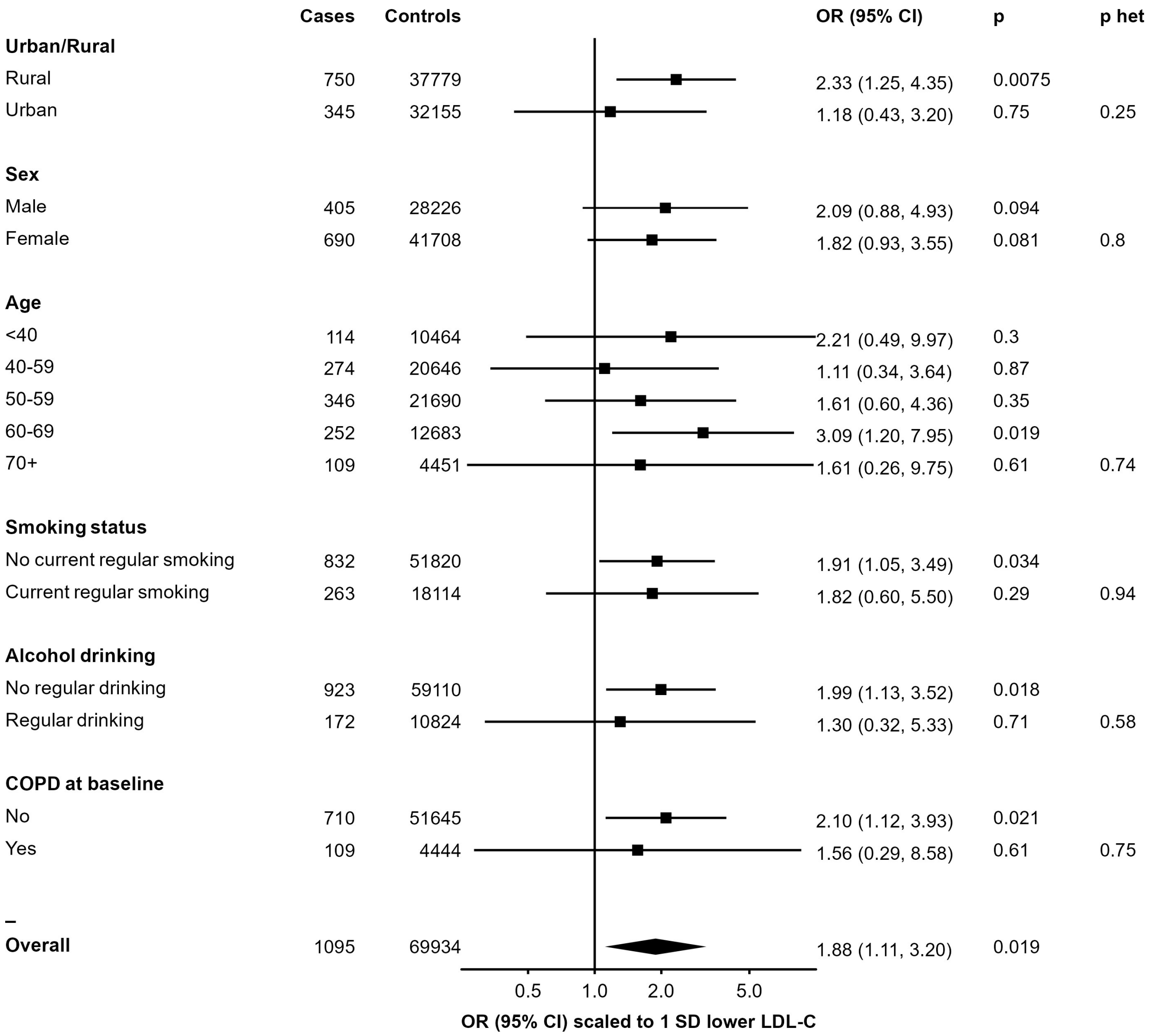

Analysis of prevalent COPD limited to 8 geographical regions in which spirometry passed quality control.
